# Metatranscriptomics reveals potential pathogens, widespread antimicrobial resistance and reduced oral microbiome diversity in acutely febrile individuals from Senegal

**DOI:** 10.64898/2026.04.16.26351042

**Authors:** Mouhamad Sy, Tolla Ndiaye, Ritika Thakur, Amy Gaye, Zoe C. Levine, Bassirou Ngom, Karina L. Bellavia, David Firer, Mariama Toure, Ibrahima M. Ndiaye, Younouss Diedhiou, Amadou M. Mbaye, Jules F. Gomis, Katherine C. DeRuff, Awa B. Deme, Mouhamadou Ndiaye, Aida S. Badiane, Marietou Faye Paye, Pardis C. Sabeti, Daouda Ndiaye, Katherine J. Siddle

## Abstract

Microbial infections elicit a range of clinical outcomes and overlapping symptoms, like acute fever, complicate diagnosis. Infecting microbes interact with resident microbiome communities, potentially driving dysbiosis and exchanging genetic material. There is thus a need to understand both the microbes associated with disease and the broader microbial context. To investigate these dynamics, we used metatranscriptomic sequencing of oral and plasma samples from febrile individuals in Senegal. Known pathogens were detected in 35% of individuals, with *Borrelia crocidurae* the most common. Viruses were present in oral (10/72) and plasma (09/78) samples, including the recently described species redondovirus, and 8 individuals had viral-bacterial co-infections. Fever was associated with reduced oral microbiome diversity and richness. Finally, a high frequency of samples expressed known virulence (24%) and resistance (10%) genes. This study sheds light on the oral microbiome in Senegal and demonstrates the utility of metatranscriptomics for integrated pathogen and resistance surveillance.

## Introduction

The human body is continually exposed to a diverse array of microorganisms—bacteria, fungi, and viruses—both from the environment and the rich communities that make up our microbiota (1–4). The outcome of these exposures is the result of an interaction between host and microbial factors (5). Microbial factors that contribute to pathogenesis, especially determinants of virulence and drug resistance, are dynamic and can be rapidly acquired and spread (6), including among microbes that establish commensal relationships with the host as part of gut or oral microbiomes (7–9). Moreover, changes to the homeostasis of these microbiome communities, known as dysbiosis, have been observed following infection (10–12), and are associated with diverse pathologies from inflammatory bowel disease (13) to pneumonia (14, 15). Together this creates a dynamic microbial landscape that is closely intertwined with health and disease.

Metagenomic and metatranscriptomic sequencing has greatly contributed to our understanding of the genomic diversity of pathogen and commensal microbes, however their application has largely been focused on the global north. Since the application of clinical metagenomics to determine the causative agent of neuroleptospirosis (16), numerous studies have demonstrated the importance of this approach for clinical diagnostics (17), outbreak investigation (18) and surveillance of emerging drug resistance (19). Studies taking a more global perspective on the gut microbiome, have identified impacts of variables including diet and industrialization on microbial prevalence and rates of Horizontal Gene Transfer (HGT) (20), a common mechanism by which bacteria can acquire resistance and virulence-associated genes. However, we lack a comprehensive understanding of these microbial communities, their composition and variability across populations and in health and disease, especially for the oral microbiome (15). Application of these techniques in understudied areas—where diagnostic testing is often more limited and where disease burdens are highest—holds potential for both improved identification of the causes of disease, and a deeper understanding of the full diversity of microbial taxa, including the prevalence of antimicrobial resistance (AMR).

In West Africa, unbiased genomic approaches have been used principally to identify pathogens associated with disease, in particular acute febrile illness (AFI) (21–24). Due to its non-specific symptoms and diverse etiologies—both infectious and non-infectious—diagnosis of AFI remains challenging (25). Tools such as metagenomic and metatranscriptomic sequencing can overcome the limited availability of diagnostic assays for surveillance (26), however, these studies generally focus narrowly on the identification of putatively pathogenic microbes, mostly viruses, in the blood. These studies often detect pathogens in only a small portion of cases and, although tnon-pathogenic species are sometimes identified, their genomic diversity, including the genes they carry, is rarely explored. Additionally, by focusing narrowly on one compartment, these studies may overlook microbial changes elsewhere in the body that are associated with disease.

Here we investigate 150 oral and plasma samples from undiagnosed acute febrile illness patients using unbiased metatranscriptomics, complemented by target enrichment and complete 16S sequencing. Plasma had low microbial diversity, with the exception of the bacterial pathogen *Borrelia crocidurae* and Human Pegivirus, which are common. We describe a complex microbiota in the oral cavity characterized by common bacterial genera—including *Capnocytophaga, Haemophilus*, *Neisseria*, and *Porphyromonas*—with infrequent presence of viruses including SARS-CoV-2 and redondovirus. We further observed a significant decrease in oral microbiome diversity in individuals with high fever, but this change was not specific to the presence of a viral infection. Finally we find that commensal oral bacteria commonly expressed genes associated with enabling host infection and antibiotic resistance, particularly beta-lactamase resistance genes. These findings help to understand pathogens and microbiome composition in the context of febrile illness.

## Results

### Population and study site characteristics

We enrolled a total of 200 individuals with a recent history of non-malarial fever across two years (2021–2022) from Linguere and Tambacounda, two climatically and demographically distinct regions of Senegal (**Figure 1A**). Tambacounda is the capital of Senegal’s largest region, Tambacounda, which is located in the southeast of the country. It has a hot, arid climate and the economy is dominated by agriculture where 93.70% of the population are farmers (27). Linguere, located in the region of Louga, is characterized by the pastoral activities of the population, many of whom have a nomadic lifestyle (27). In both localities, the majority of participants were female, 56.56% in Linguere (n = 56/99) and 68.31 % in Tambacounda (n = 69 /101) (**Figure 1B**), compared to 49.3% in the general population in 2024 (27). The ages of participants ranged from 2 years to 85 years (**Figure 1B**), with a median age of 19 years, similar to that of the Senegalese population as a whole (27, 28). Participants from Tambacounda were, on average, younger than those from Linguere (**Figure 1B**). While all participants enrolled in this study had a recent history of fever (in the last three days), underarm temperature taken upon enrollment ranged from 36 - 39 °C, and the median temperature was 38 °C (**Figure 1B**).

**Figure 1:**
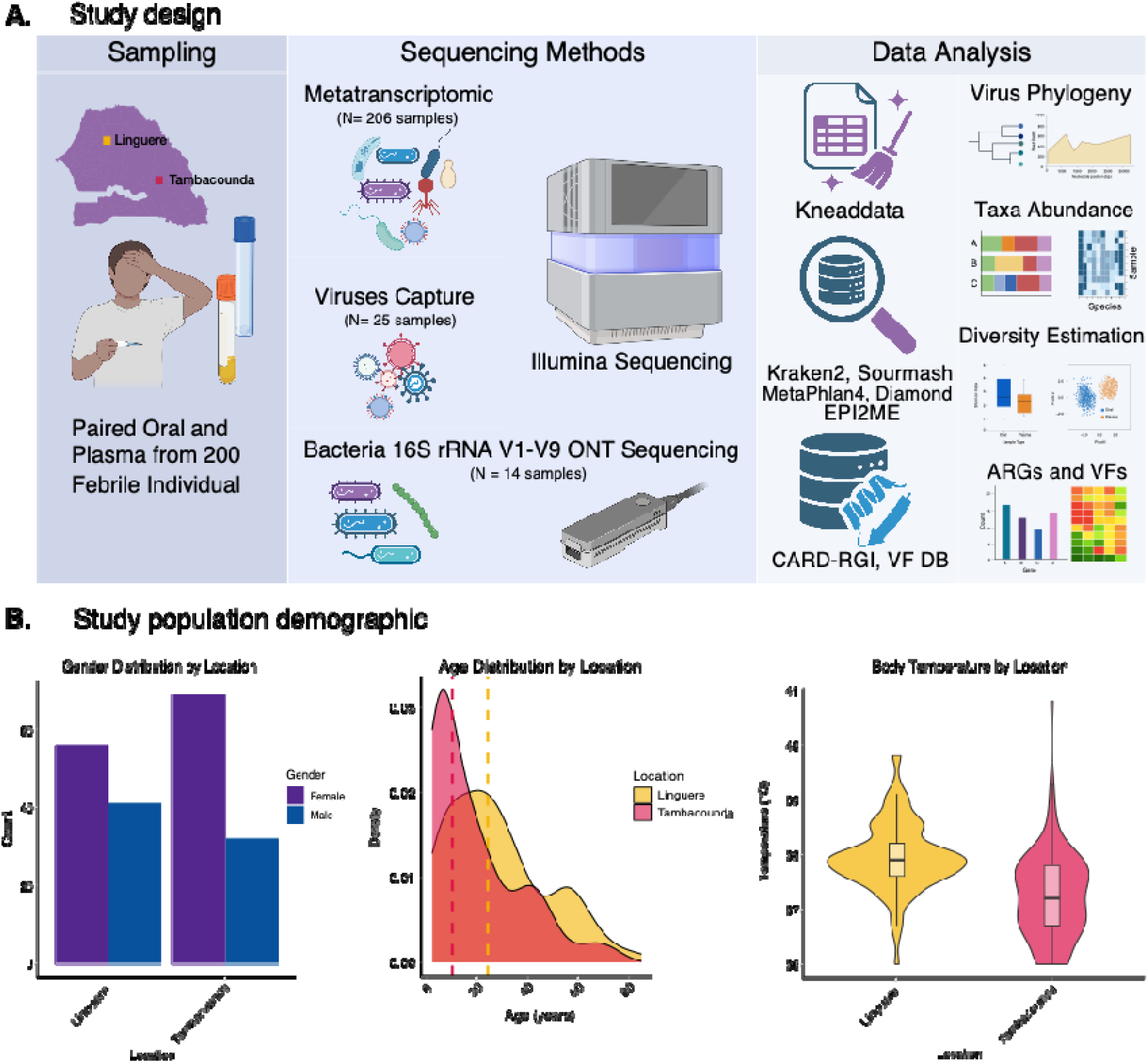
Study overview. A) Study design showing sampling strategy, all sequencing methods used and a simplified data analysis workflow. Panel A figures were created with Biorender B) Study population demographics with gender, age and body temperature distribution across Linguere and Tambacounda.

### Microbial communities and diversity in plasma and oral samples

After filtering for library quality and sequencing depth (>700,000 paired-end reads), a final dataset of 1,455,296,788 paired-end reads were obtained from 150 samples (72 oral and 78 plasma). The mean and median paired-end read counts were 9,701,979 and 3,992,135, respectively [714,698 – 140,414,520] and there was no difference in sequencing depth between sample types (**Supplementary Figure 1**). To reduce the risks of misclassification or overestimation of microbial taxa, we removed human reads using the kneadData v0.12.3 pipeline (29) and further filtered the results for; i) potential contaminations using Decontam (30) a list of likely experimental and environmental contaminants (**Methods & Supplementary Table 1**), and ii) taxa with fewer than 100 reads (count). As previous studies have shown that individual metagenomic classifiers are prone to errors in classification (31), we used two different tools: Kraken2 (a read based classifier) (32), sourmash v4.9.4 (a k-mer based classifier) (33). Kraken2 and sourmash showed a strong correlation in the taxa they identified and their abundance estimates (**Supplementary Figure 2**), and rarefaction curves indicated sufficient sequencing depth to capture communities present in each sample type (**Supplementary Figure 1**).

To characterize the microbial communities present in plasma and oral samples, we first evaluated their taxonomic composition and diversity. As expected, bacteria were the most abundant domain and were significantly more abundant in oral samples compared to plasma (p<10e-16; **Figure 2A**). In oral samples, bacterial reads were dominated by common members of the oral microbiome (34) including *Capnocytophaga, Haemophilus*, *Neisseria*, *Porphyromonas*, *Prevotella*, *Pasteurella*, and *Veillonella.* Abundance estimates were broadly similar, although some genera were only identified in one of the two methods (**Figure 2B**). In plasma, the most frequent and abundant genera reflect either, i) contamination from the skin microbiome, as in the case of *Cutibacterium*, the most abundant genus in plasma samples (35), or ii) translocation from oral or gut microbiomes, such as *Porphyromonas* and *Prevotella*, as described in other studies (36, 37). One exception is *Borrelia* (*B*.), a spirochete with several pathogenic species. There was considerable inter-individual variability in metatranscriptomic profiles, especially in plasma (**Supplementary Figure 3**), consistent with the low biomass of this sample. Despite the overlap in certain common taxa, oral and plasma samples showed clearly distinct microbial profiles, with Bray–Curtis distance–based PCoA showing distinct clustering of the two compartments and a significant difference by PERMANOVA (R² = 0.14, p = 0.001) (**Figure 2C**). Using Kraken2 counts, oral samples showed higher Shannon Index (mean Shannon = 3.50 ± 0.59; n = 72) compared to plasma samples (mean Shannon = 0.89 ± 1.28; n = 78) suggesting low biomass in plasma (**Figure 2D**).

**Figure 2:**
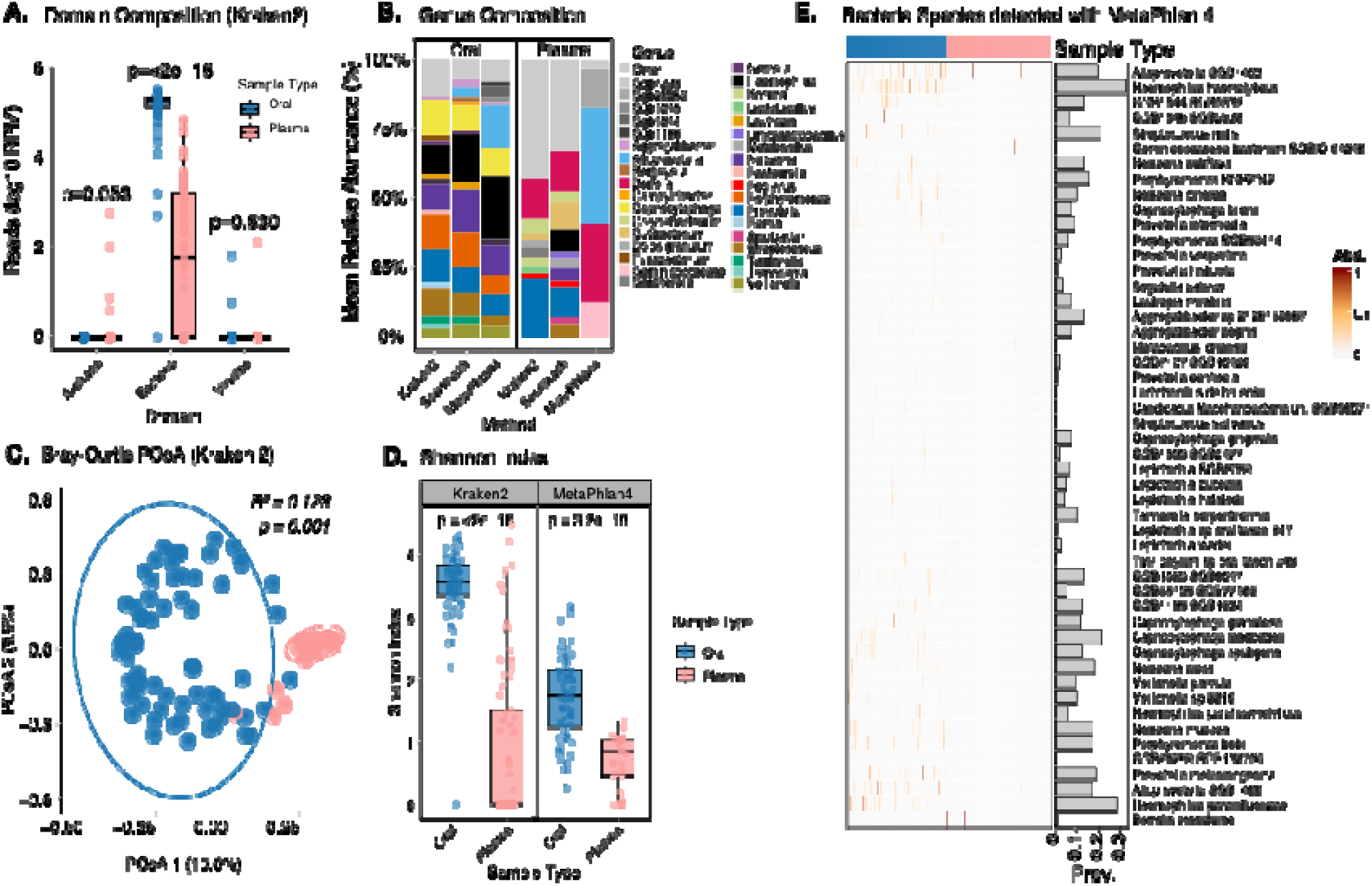
Taxonomic composition and diversity. A) The log10 of total read counts per million (RPM) for the four detected domains Archaea, Bacteria and Viruses in both oral sample (blue) and plasma sample (coral). A Wilcoxon rank-sum test is performed for each domain comparison, with the resulting p-value displayed at the top to indicate statistical significance in total abundance. B) mean relative abundance of genera in Oral and Plasma detected by the tree methods. C) Principal Component Analysis (PCoA) Bray-Curtis dissimilarity beta diversity for oral and plasma. Percentage in axis 1 and 2 represent the proportion of variance. The R^2^ and p-values at the top are derived from PERMANOVA. D) Alpha diversity Shannon index, p value represents the Wilcoxon rank-sum test at the species level estimated with Kraken2 and MetaPhlan4. E) abundance (heat map) and prevalence (vertical barplot) of the top 50 species detected with MetaPhlan4 by sample type.

### Resolving bacterial presence at the species level

We next searched for potential pathogens that could be associated with the AFI of these individuals. As the sequence similarity between common pathogenic and non-pathogenic species can make resolving bacterial identities difficult from short read sequencing data, we incorporated here the results of MetaPhlan4 (38), a marker-based method that is known to have lower sensitivity but higher specificity than k-mer based methods such as Kraken2 (39, 40). MetaPhlan4 recapitulated previously observed trends in genus-level taxa and sample richness and diversity (**Figure 2B & Figure 2D**). In total 163 unique bacterial species were identified with MetaPhlan4 (**Figure 2E)**. In plasma, *B*. *crocidurae* was identified in two samples from Linguere (**Figure 2E**). Kraken2 and sourmash indicated 2 additional samples, also from Linguere, with *B*. *crocidurae* and sourmash alone suggested *B. crocidurae* in a single sample from Tambacounda (**Supplementary Figure 4**). *Borrelia*, particularly the species *B. crocidurae*, is known as one of the main causes of relapsing fever in Senegal (41). Very few other taxa were detected in plasma, consistent with the absence of a blood microbiome (35). Taxa that were detected, e.g., *Lactobacillus delbrueckii, Cutibacterium acnes, Alloprevotella SGB1463,* and *Staphylococcus epidermidis,* are suspected to be present in blood either by translocation from areas like the gut or mouth (42), or contamination from the skin during venipuncture. In oral samples, MetaPhlan4 detected only common oral microbiota species including *Prevotella melaninogenica, Capnocytophaga leadbetteri, Tannerella serpentiformis, Streptococcus mitis, Haemophilus haemolyticus, Haemophilus parainfluenzae, Neisseria mucosa, and Neisseria cinera* (**Figure 2E**) (43–45).

To complement our metatranscriptomic data and further inform species-level identities in oral samples, we performed total 16S rRNA (V1-V9) sequencing using Oxford Nanopore Technology (ONT) on 9 oral samples. A recent study has shown that ONT has a lower error rate with the new chemistry and provides better accuracy for bacterial species identification using full-length 16S rRNA (46). After data cleaning and trimming we obtained an average of 153,200 reads between 1.2 and 1.8 kb per sample. Rarefaction confirmed that sequencing depth was sufficient in all samples (**Figure 3A**). The majority of species detected in metatranscriptomics were replicated in 16S rRNA **(Figure 3C** and **Supplementary Figure 5**). For abundance estimates, MetaPhlan4 showed the highest correlation with 16S sequencing at the species level, supporting that this was the most appropriate tool for interpreting metatranscriptomic sequencing results (**Figure 3D**). However, we observed differences in abundance estimates for certain species. *Neisseria oralis, Streptococcus oralis,* and *Streptococcus gordonii* showed consistently higher abundance in 16S compared to metatranscriptomics (**Figure 3E)**, suggesting differences between species presence and transcriptional activity, as previously observed for other bacteria (47, 48). Other species showed more variability in abundance levels between methods (**Supplementary Figure 6)**, and we also observed variability between samples in the concordance of abundance estimates (**Supplementary Figure 7).** Together, these results highlight some of the challenges of resolving sequencing data to the species level and broadly support the absence of major bacterial respiratory pathogens in this cohort.

**Figure 3:**
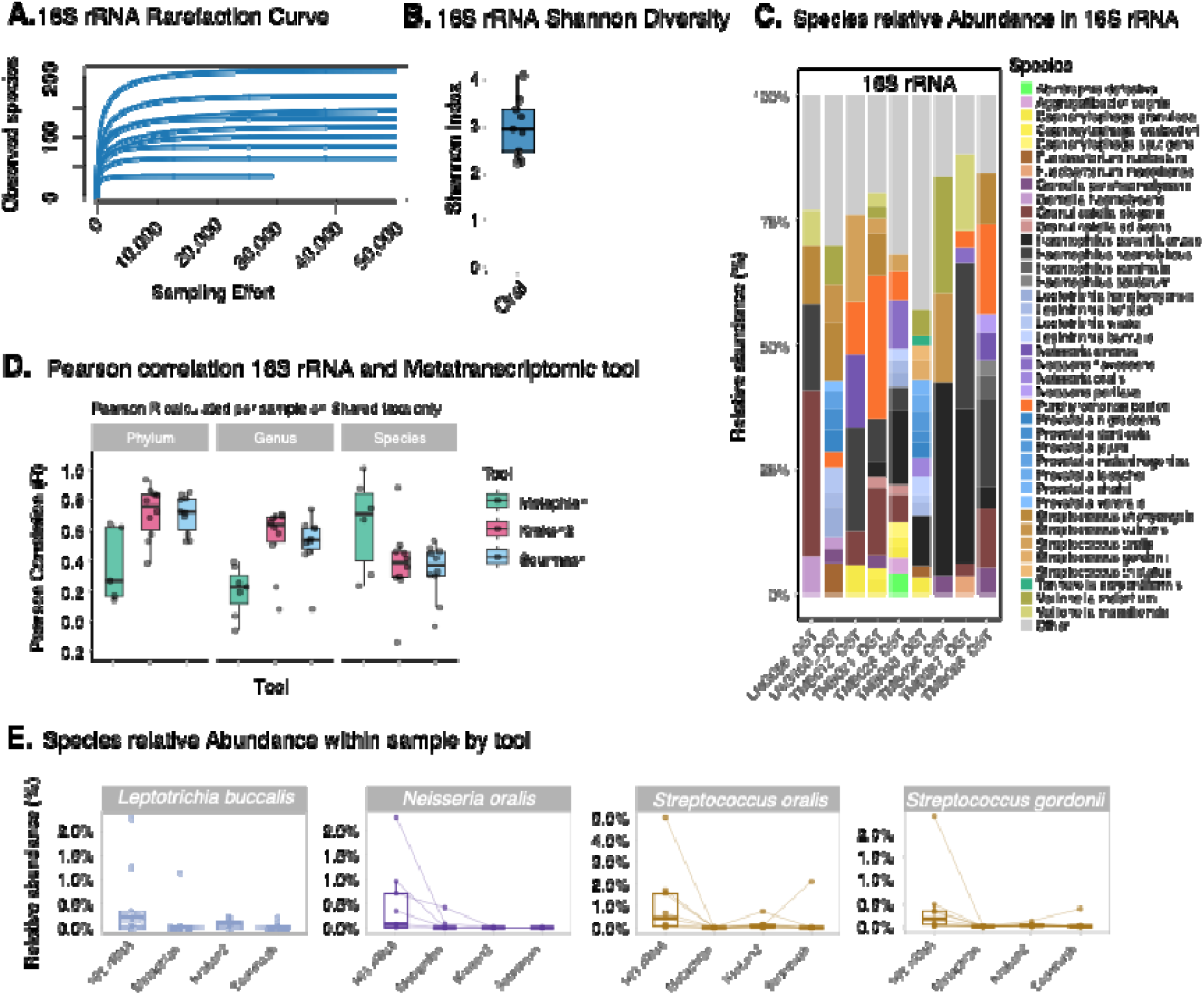
Total 16S rRNA and metranstranscriptomic concordance analysis. A) rarefaction curve at the species level. B) Shannon diversity index. C) species relative abundance. The category “other” in light grey represents species with less than 1% abundance. D) Pearson correlation between MetaPhlan, Kraken2 and sourmash compared to 16S at the Phylum, Genus and Species rank. E) relative abundance of species for each tool. Here, each dot corresponds to a single sample; connecting lines are used to visualize how abundance changes for a given sample depending on the tool used.

### Infrequent detection of viral pathogens in plasma and oral samples

We further screened metatranscriptomic sequencing data for the presence of RNA and DNA viruses. We identified human-infecting viral genera in 19 patient samples (9 plasma and 10 oral samples), using either Kraken2 or Sourmash (**Methods** and **Figure 4A**). The most frequently detected viral genus was betacoronaviruses (SARS-CoV-2) found in 5 oral samples, 3 from Linguere and 2 from Tambacounda. Torbevirus and pegivirus were detected in 5 oral and 3 plasma samples, respectively. Four of these viruses —betacoronavirus, lymphocryptovirus (species: Epstein Barr Virus), Mastadenovirus (species: Human mastadenovirus C) and Simplexvirus (species: Human alphaherpesvirus 1)—are known human pathogens. Pegivirus, commonly detected in all human populations, is believed to be non-pathogenic (49). As viral sequences, especially in poorly described taxa or regions, may be highly divergent from those in the databases used by taxonomic classification tools, we further used the protein-based tool DIAMOND to classify contigs assembled with SPAdes (see Methods and **Supplemental Table 3**). High-identity matches in DIAMOND (>90%) confirmed several of the previously detected hits for Betacoronavirus, Simplexvirus, Pegivirus, Torbevirus (unclassified Redondoviridae), and Alphatorquevirus, and identified 1 additional Pegivirus positive sample (**Figure 4B**).

**Figure 4:**
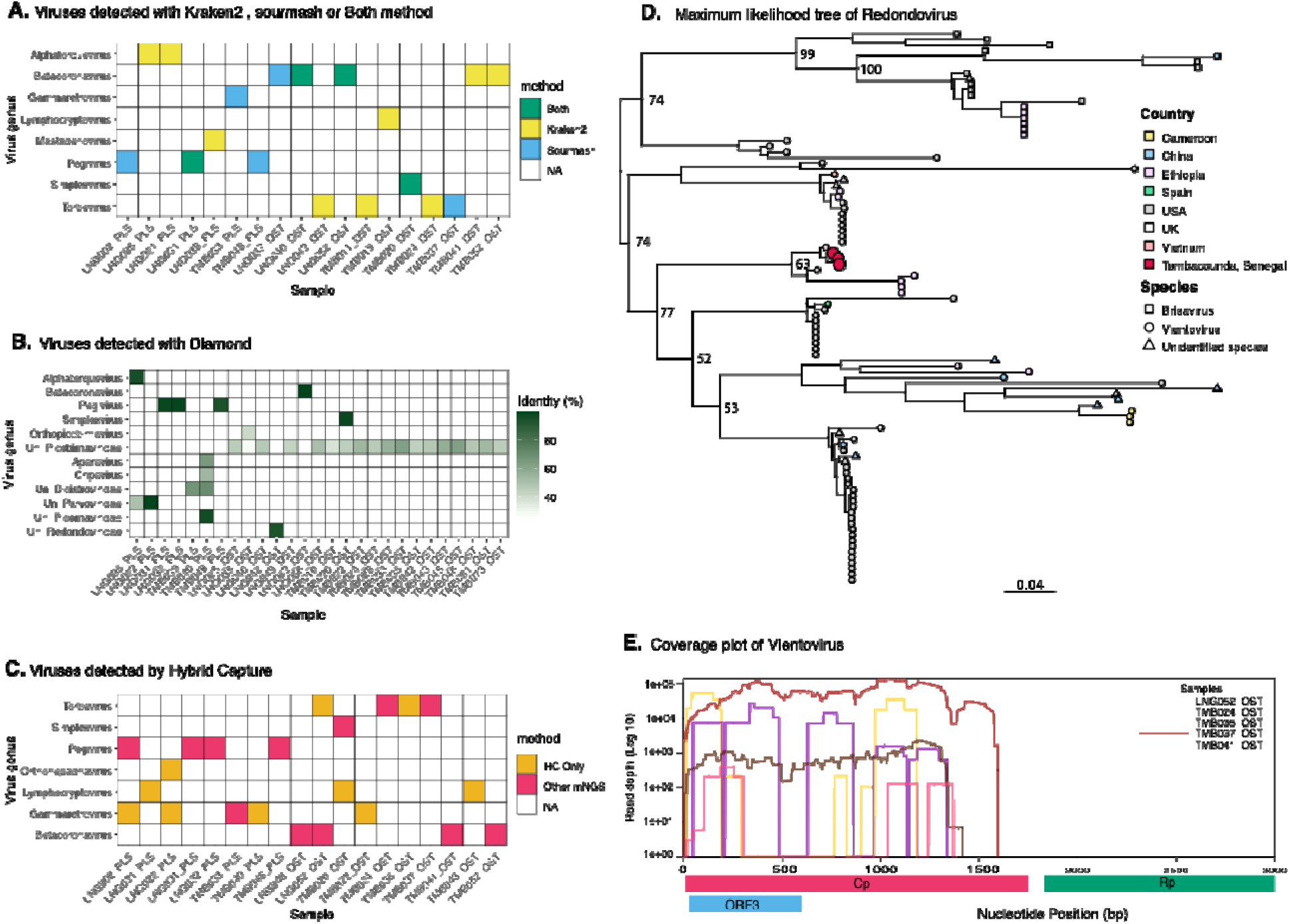
Viral detection in oral and plasma samples. A) Viral genera detected by classifiers Kraken2 and sourmash, overlap between tools is shown in green. B) Viral genera detected by DIAMOND. Colour depth reflects percent identity of the contig to know sequences. Genera labels beginning with “un_” unclassified members of this group. C) Viral genera detected by hybrid capture showing both confirmation of findings from unbiased sequencing and new taxa detected following enrichment. D) Maximum likelihood tree of whole redondovirus genomes. Node shape denotes species and colour denotes origin. Larger nodes are new samples from this study. E) Coverage plot of vientovirus genomes showing deep coverage of the capsid gene in samples from this study.

Among more divergent sequences (those with 20-80% identity or where contigs could only be confidently classified to the Family level), DIAMOND identified 2 plasma samples with contigs matching to the family Parvoviridae, and 2 other plasma samples with contigs of the family Dicistroviridae (**Figure 4B)**. Dicistroviridae is a recent virus described in human febrile patients, as reported in recent studies conducted in Nigeria (50), Tanzania (51), and Peru (52). One of these samples (TMB040_PLS) also contained a contig matching (with 96.9% identity) to the family Picornaviridae. To improve our recovery of viral genomes, we performed pan-viral enrichment using TWIST’s comprehensive viral panel on a total of 25 samples, of which 60% (15/25) were oral samples and 40% (10/25) were plasma. We recovered an average of 10.7 million reads per sample (**Supplementary Figure 8**). After enrichment, we were able to recover complete or partial genomes for SARS-CoV-2 (n=3), Epstein-Barr Virus (n=1), Human Pegivirus C (n=3) and Redondovirus (n=5) (**Figure 4C** and **Supplementary Tables 4**).

Redondovirus, a recently described circular single stranded DNA virus of the respiratory tract (53, 54), was the most frequent virus detected in our cohort. We successfully assembled the majority of the capsid gene (positions 1–1593 of the genome) in 3 samples from Tambacounda. Two additional samples (1 from Linguere and 1 from Tambacounda) assembled only a few hundred bases of the capsid gene. We performed phylogenetic analyses comparing our sequences to publicly available Rendoviridae genomes to contextualize these findings within existing viral diversity. Whole-genome phylogenetic analysis including the 3 more complete sequences showed that our samples are the species vientovirus, which clusters distinctly from brisavirus, the only other species in this viral family (**Figure 4D**). Within the vientovirus clade, these 3 sequences (all from Tambacounda) clustered together most closely related to sequences from the United States, though this likely reflects unequal sampling of redondoviruses which have mostly been sequenced in samples from the U.S.. We suspect that we were only able to assemble the capsid gene sequence for these samples (**Figure 4E**) due to the lack of complementary probes to the more genetically diverse Rep gene in the capture panel. Phylogenies of the capsid gene alone, and of the 412 base pairs of sequence assembled for all 5 samples (**Supplementary Figure 9**), show the sequence from Linguere is genetically distinct from those from Tambacounda, suggesting geographic structuring of viral diversity, however, more complete genomes are needed to confirm this. These samples were also from different years, however, a root-to-tip plot shows no strong temporal signal in the tree (**Supplementary Figure 9**). Crucially, since no other genomes of this species are available from West Africa making inference challenging and highlighting the importance of viral surveillance in understudied regions to fully dissect viral diversity.

### Fever is associated with a reduction in oral microbiome diversity

As fever has previously been associated with dysbiosis in the gut microbiome (55), we next investigated community-level changes in the oral microbiome associated with disease. Linear regression identified a significantly lower species richness and diversity (p=0.04) in individuals from Linguere with elevated body temperatures (**Figure 5A**). As no individuals from Tambacounda had an acute fever (defined as a temperature >38°C) at the time of enrollment, we cannot evaluate further in that population. We see no differences in diversity and richness associated with sex, age or location (**Figure 5A**). However, community composition differed between Linguere and Tambacounda (**Figure 5B**). Looking at changes in specific taxa with temperature, we found that the genus *Haemophilus*, and in particular the species *Haemophilus haemolyticus*, showed substantially higher relative abundance in individuals with acute fever (**Figure 5C**).

**Figure 5:**
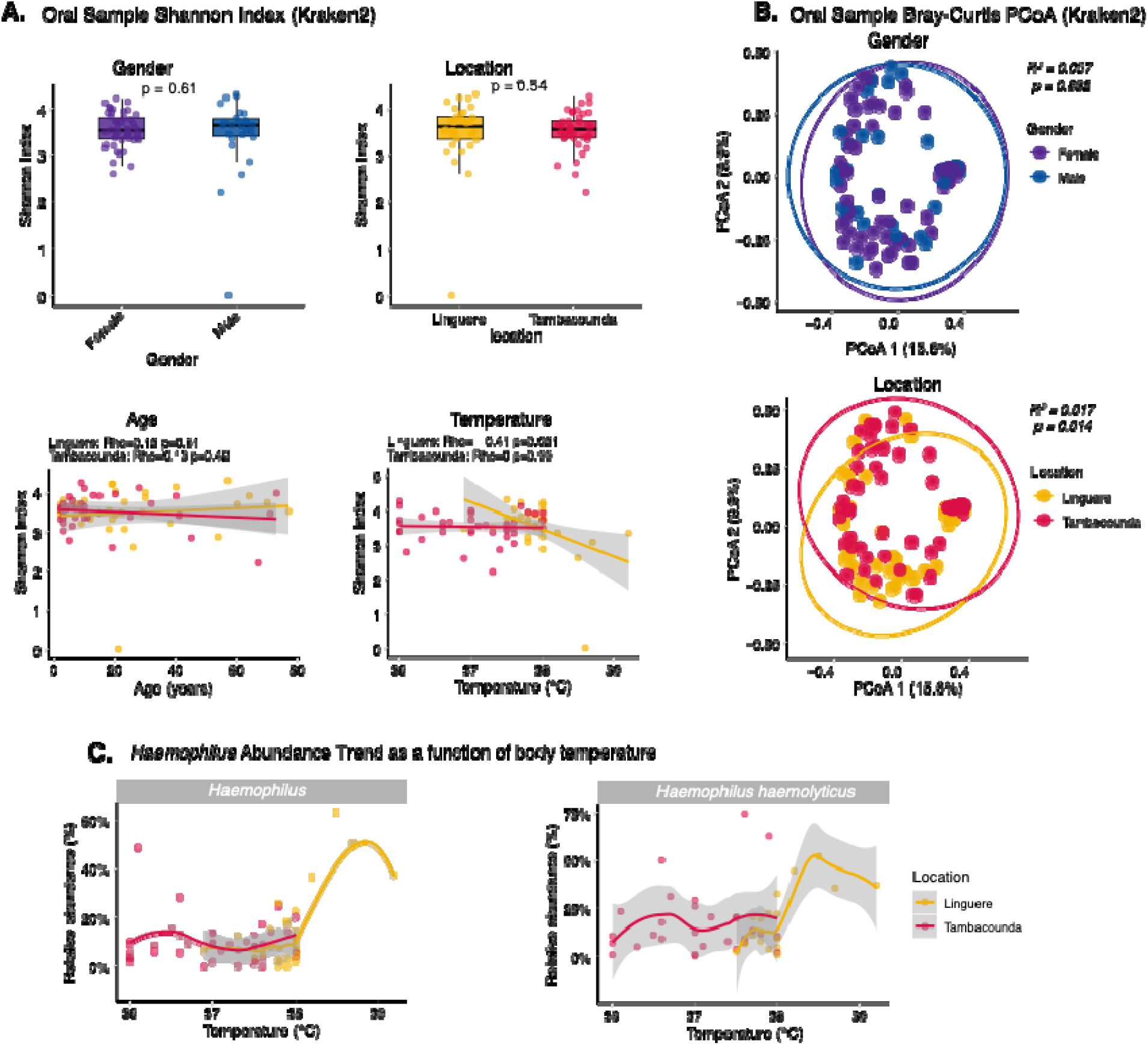
Changes in taxonomic diversity in oral samples. A) Shannon diversity index for oral samples across age, gender, location, and temperature. P-values represent Wilcoxon rank-sum test results. B) Principal Coordinates Analysis (PCoA) of Bray-Curtis distance, illustrating the similarity of oral microbial composition by gender and location. The R^2^ and p-values at the top are derived from PERMANOVA; no significant differences were observed for either gender or location (p > 0.05). Each 95% concentration ellipse estimates the region where 95% of the samples are expected to cluster. Percentage in axis 1 and 2 represent the proportion of variance. C) Scatter plots showing relative abundance of the genus *Haemophilus* and the species *Haemophilus haemolyticus* compared to measured body temperature for Linguere and Tambacounda. The solid lines represent trends for each location.

We also considered whether presence of specific viral taxa was associated with differences in the oral microbiome. Overall, there was no significant difference in Shannon Index between species where at least one virus was detected and those where no viral taxa were detected (**Supplementary Figure 10**). For respiratory viruses SARS-CoV-2 and Vientovirus we again found no difference between individuals with and without these respiratory infections (**Supplementary Figure 10**), suggesting that viral presence is not associated with species richness or diversity.

### Haemophilus and Neisseria exhibit high frequency of genes linked to resistance and virulence

The microbiota serves as a complex reservoir for AMR (56), where commensal bacteria can protect pathogens by degrading antibiotics (7) and facilitating HGT, as shown for beta-lactam resistance in the human gut (57), highlighting the critical importance of understanding antimicrobial resistance genes (ARG) within these environments. We used metatranscriptomic contigs >500bp assembled from our 150 samples with MEGAHIT v1.2.9 (58), to identify ARGs with CARD-RGI (see Methods). ARGs were detected in 19 samples and in 15 unique bacteria species **(Figure 6A, Supplementary Table 5**). In total, 17 distinct ARGs were identified (**Figure 6A**), belonging to seven major antimicrobial drug classes of which beta-lactam resistance genes were the most common (n=15) (**Figure 6A**). These ARGs spanned several canonical resistance mechanisms; enzymatic inactivation was the most frequent (n=14), followed by target alteration (n=10) (**Figure 6A**). The most common single ARG was *Ecol_EFTu_PLV*, present in 9 oral samples (12.6%). This gene, which confers resistance to pulvomycin (59), was consistently associated with *Haemophilus* species by BLAST identity search (**Figure 6B**). geNomad indicated this gene to be primarily chromosomal with some disagreement between samples (**Table 1**). Other common gene families were the *TEM* and *CfxA* families of beta-lactamase enzymes (60). *CfxA* genes were associated with *Bacteroides* and *Prevotella* (**Figure 6B**). As TEM genes were predominantly located on mobile elements (plasmid scores > 0.90), the species assignments for these may be less accurate (**Table 1**).

**Figure 6:**
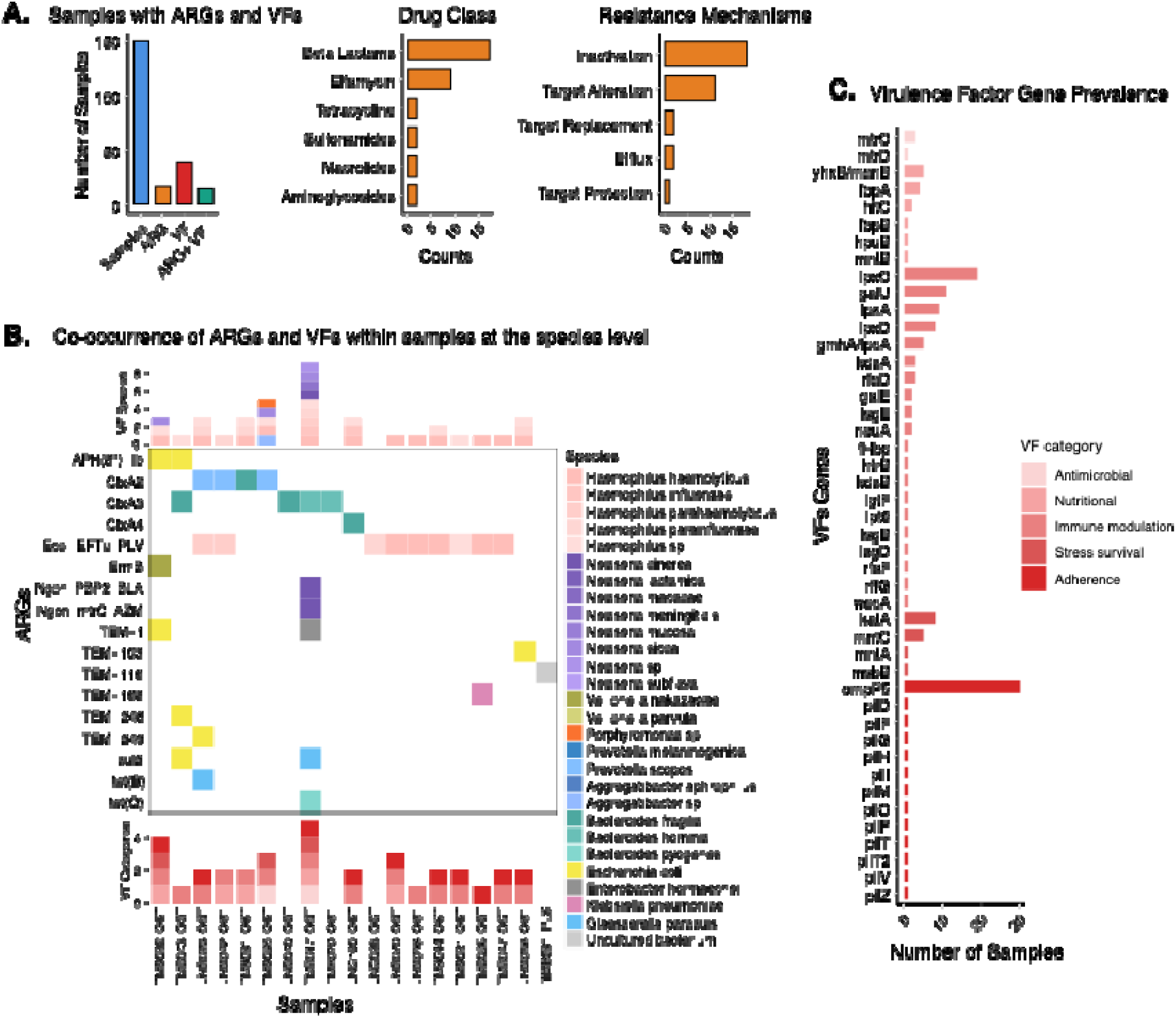
Antimicrobial resistance (ARG) and virulence factors (VF) genes identified across species. A) summary of samples with AMR genes and virulence factors, ARG drug class and resistance mechanisms counts. B) ARG heatmap highlighting bacterial species associated with specific AMR gene families and virulence factors in samples where both are detected. C) prevalence of virulence factor genes across samples, grouped and color-coded according to their biological categories (antimicrobial, immune modulation, adherence, stress survival, nutritional).

**Table 1:** ARGs origins predicted based on geNomad contig aggregated classification score.

| ARG | Aggregated classification score |  |  | Predicted origin | Sample |
| --- | --- | --- | --- | --- | --- |
|  | Chromosome | Plasmid | Virus |  |  |
| *APH(3'')-Ib* | 0.366 | **0.441** | 0.192 | Plasmid* | TMB022_OST |
|  | 0.336 | 0.280 | **0.385** | Virus* | TMB043_OST |
| *cfxA2* | 0.154 | 0.144 | **0.702** | Virus | LNG023_OST |
|  | 0.194 | 0.224 | **0.582** | Virus | TMB031_OST |
|  | 0.276 | 0.269 | **0.456** | Virus* | LNG042_OST |
|  | 0.271 | 0.340 | **0.389** | Virus* | TMB035_OST |
| *cfxA3* | 0.194 | 0.184 | **0.622** | Virus | TMB020_OST |
|  | 0.261 | **0.404** | 0.334 | Plasmid* | LNG040_OST |
|  | 0.265 | 0.341 | **0.394** | Virus* | TMB017_OST |
| *cfxA3 / TEM-248* | 0.003 | **0.987** | 0.009 | Plasmid | TMB043_OST |
| *cfxA4* | 0.209 | **0.663** | 0.128 | Plasmid | LNG100_OST |
| *Ecol_EFTu_PLV* | 0.023 | **0.961** | 0.016 | Plasmid | TMB036_OST |
|  | **0.952** | 0.029 | 0.019 | Chromosome | LNG070_OST |
|  | **0.935** | 0.038 | 0.027 | Chromosome | LNG098_OST |
|  | **0.724** | 0.141 | 0.135 | Chromosome | TMB047_OST |
|  | 0.121 | 0.178 | **0.702** | Virus | LNG042_OST |
|  | **0.660** | 0.163 | 0.177 | Chromosome | TMB021_OST |
|  | **0.637** | 0.158 | 0.205 | Chromosome | LNG023_OST |
|  | **0.601** | 0.170 | 0.230 | Chromosome | TMB014_OST |
|  | 0.278 | 0.163 | **0.559** | Virus | LNG028_OST |
| *ErmB* | 0.192 | 0.308 | **0.500** | Virus | TMB022_OST |
| *NG_mtrC* | **0.764** | 0.207 | 0.029 | Chromosome | TMB017_OST |
| *NG_PBP2* | **0.967** | 0.024 | 0.010 | Chromosome | TMB017_OST |
| *sul2* | 0.075 | **0.824** | 0.101 | Plasmid | TMB043_OST |
|  | 0.174 | **0.562** | 0.264 | Plasmid | TMB017_OST |
| *TEM-1* | 0.003 | **0.988** | 0.009 | Plasmid | TMB022_OST |
|  | 0.004 | **0.983** | 0.012 | Plasmid | TMB017_OST |
| *TEM-102* | 0.026 | **0.926** | 0.048 | Plasmid | LNG056_OST |
| *TEM-116* | 0.010 | **0.963** | 0.027 | Plasmid | TMB031_PLS |
| *TEM-163* | **0.679** | 0.155 | 0.167 | Chromosome | TMB036_OST |
| *TEM-249* | 0.007 | **0.989** | 0.004 | Plasmid | LNG023_OST |
| *tet(B)* | **0.614** | 0.223 | 0.163 | Chromosome | LNG023_OST |
| *tet(Q)* | 0.172 | **0.655** | 0.173 | Plasmid | TMB017_OST |

Scores are geNomad marker- and neural-network-derived probabilities aggregated across the contig carrying the ARG; the three scores sum to 1 for each contig. The higher score are in bold and the asterisk marks predicted origin which score <0.50.

Virulence Factors (VFs) are proteins that enable a bacteria to cause disease, by contributing to processes such as adhesion, invasion, toxin production, immune evasion, or survival within the host (61, 62). To assess the presence of genes encoding known VFs, we screened all our assembled samples against the Virulence Factor Database (63). Out of the 150 samples, 37 samples (24%) harbor at least one VF gene across five categories: adherence, antimicrobial, immune modulation, nutritional, and stress survival. VFs were detected in 18 unique bacteria species (**Figure 6C, Supplementary Figure 11**). The adherence-associated gene *ompP5*, was the most prevalent gene, appearing in nearly 30 samples and associated with *Haemophilus*, *Prevotella* and *Veillonella* species (**Supplementary Figure 11**). Several immune-modulation genes, such as *lpxC*, *galU*, *lpxA*, *lpxD*, and *gmhA*/*lpcA*, showed moderate prevalence and were again mostly present in *Haemophilus* species alongside *Aggregatibacter* (**Supplementary Figure 11**). Nutritional acquisition genes (*yhxB*/*manB*, *fbpA*, *hitC*) and stress-survival factors (katA, mntC, msbB, mntA) appeared in a small set of samples and were mostly associated with *Haemophilus* and *Neisseria* species. This approach also identified antimicrobial-related genes, confirming the presence of *mtrC* in *Neisseria* and also detecting a second gene, *mtrD*.

As the co-existence of ARGs and VFs can make bacteria resistant to treatment and cause severe disease (64), we finally evaluated overlap in our cohort. While 15 samples showed co-occurrence of ARGs and VFs, *Haemophilus* and *Neisseria* were the only species found to harbor both genetic elements simultaneously (**Figure 6B**). This is most clearly illustrated by *TMB017*, which was sequenced to very high depth (140 million paired end reads). In this sample we detected 6 ARGs, including two chromosomally present resistance mutations in *Neisseria cinera* (*NG_PBP2, NG_mtrC*). This sample also contained a cluster of 11 pilus-associated adherence genes (*pilZ*, *pilV*, *pilT*, *pilP*, *pilO*, *pilM*, pilI, *pilH*, *pilG*, *pilF*, *pilD*)(65), also assigned to this species (**Supplementary Figure 11**). Given the high sequencing depth of this sample, we suspect that our findings are likely not exhaustive of all virulence-associated genes present in our cohort of samples and deeper sequencing would detect more such overlaps. Consistent with this, a co-occurrence was also identified in *Prevotella scopos* in sample TMB035, where we detected both *cfxA2* and species-specific virulence genes.

## Discussion

In this study, we combined complementary sequencing approaches in a cohort of paired oral and plasma samples with AFI to investigate potential underlying pathogens and the circulation of ARGs in Senegal. This study provides a broad characterization of the oral microbiome from Senegal—across 72 individuals from two distinct regions—and identifies a reduction in diversity and richness associated with fever. The oral microbiome is important because it plays a vital role in maintaining overall systemic health (34, 44). Previous studies have observed that the homeostasis of the gut microbiome can be modified by body temperature (66), and specific infections, such as SARS-CoV-2 are associated with changes to both the oral and gut microbiome (10–12). However, the effects of fever more broadly on the microbiota are poorly understood. Our finding that individuals with a high fever (defined as a temperature at enrollment measuring >38°C) show significantly reduced alpha diversity compared to individuals with normal or slightly elevated body temperatures suggests a more general association. We identify *Haemophilus* as the genus showing the most marked increase in relative abundance among individuals with fever in metatranscriptomic data. More extensive investigation of relative abundance of microbial DNA would reveal if these changes reflect underlying changes in microbe abundance or changes in gene expression, such as upregulation of heat-shock-related genes. Beyond bacterial taxa, we identify and assemble complete capsid gene sequences for four isolates of *Torbevirus viento*. These sequences, the only ones from West Africa to date, contribute to improving our understanding of the diversity of this recently defined species.

A key insight from this work is the high prevalence of ARGs, especially genes conferring resistance to beta-lactams, in our cohort. AMR is a pressing public health threat in Senegal and globally. Overuse of antibiotics is recognized as driving the emergence of AMR, and the untargeted use of antibiotics has been found to be most common in patients without a diagnosis (67). While only 2 individuals in our cohort had taken antibiotics for their most recent illness, beta-lactams represent one of the most widely used classes of antibiotics in Senegal, accounting for nearly 90% of antibiotic prescriptions (68). We find that 19% of oral samples (n=14/72) contain at least one known ARG for beta-lactams. Genes of both the TEM and CfxA families were detected, and ARGs were identified in both sampling locations (Linguere and Tambacounda). Previous studies in Senegal have identified significant levels of beta-lactam resistance in a wide variety of settings, including hospitalized patient samples (69, 70) and broiler chickens (70). However, all of these studies rely on targeted PCR assays focusing on known ARGs, sometimes coupled with culture-based antimicrobial susceptibility testing. Our use of metatranscriptomic sequencing offers a complementary approach providing a broad, untargeted means to identify ARGs.

We identify putative pathogens in a modest number of cases (n=23, **Supplementary Table 2**), broadly consistent with other similar studies (24), highlighting the challenges of diagnosing AFIs even with genomic tools. *Borrelia crocidurae* is the most detected pathogen (Linguere n=10 and Tambacounda n=4), consistent with our previous findings in Thies (22). We also identified cases of SARS-CoV-2 in both locations (Linguere n=3, Tambacounda n=2), while several other pathogens were detected in just one or two individuals. Some pathogens previously found to be common in acute respiratory infections (ARI) in Senegal were not identified here. One reason for this may be differences in timing: many common viral pathogens such as adenoviruses, rhinoviruses, influenza viruses, and coronaviruses are known to exhibit strong seasonality (71–73). *Streptococcus pneumoniae*, found by one study to be present in 74% of ARI in Keur Socé (74), was also not detected in this study. These differences may be attributable to differences in collection location, sample type, and/or enrollment criteria. We also note that *Streptococcus* species were more abundant in 16S sequencing than metatranscriptomics, although no *S. pneumoniae* was identified in the 9 oral samples tested by ONT sequencing. Other factors that may explain the absence of potential pathogens even in individuals with active or recent fever include non-infectious conditions such as autoimmune diseases which may also cause fever (75) and missing the period of active infection where microbial RNA/DNA can be detected. Pathogens may also be present in other body compartments: while in this study we considered both oral and blood compartments which are common reservoirs for infections, pathogens in the gastrointestinal tract or central nervous systems, for example, would not be detected.

Our study has some limitations. First, our cohort is composed exclusively of febrile individuals, and we do not have data from healthy (afebrile) individuals from the same time and locations. We therefore cannot assume that the presence of specific microbial species is the direct cause of fever, as we lack knowledge of the baseline microbial composition in the general population (76). As the main findings of our study concern the wider microbial communities, this does not significantly impact our conclusions but would be an important consideration for any further work investigating the biological significance of poorly characterized viral taxa, such as Dicistroviridae and Redondoviridae. Second, our choice of metatranscriptomic sequencing, while offering the potential to detect microbes with both DNA and RNA genomes simultaneously, means we are unable to assemble complete genomes for DNA-based organisms. This is a particular challenge for understanding the larger sequence context of ARGs and VFs, however transcript assembly did provide some insight into this, allowing us to classify 90% of our ARGs and VFs at the species level.

In conclusion, this study provides novel insights into the landscape of microbes circulating in febrile individuals in Senegal, highlighting the importance of investigating microbial communities using untargeted methodologies to explore their associations with disease. Moreover, and in light of recent emphasis on the importance of expanding surveillance of AMR in Senegal under a One Health approach (77), our results highlight the value of integrating genomic approaches with existing pathogen and AMR identification tools to leverage their capacity to detect and characterize ARGs that may not be included in routine surveillance in resource-limited settings.

## Methods

### Sample collection

Patient samples were obtained through a study approved by the Institutional Review Boards at the Senegalese Ministry of Health and Harvard University. Patient samples were obtained from febrile individuals presenting to the hospital in Linguere and Tambacouda, Senegal. Study staff obtained written, informed consent from all participants enrolled in the study (or a patient/legal guardian for participants age less than 18 years) following an explanation of the study objectives. All methods were performed in accordance with the relevant guidelines and regulations. A 5 mL venous blood draw and an oral swab into Trizol were collected from each enrolled patient. Demographic information as well as clinical signs and symptoms were also recorded. Samples were kept on liquid nitrogen and transported to Dakar for further processing.

### RNA extraction and sequencing

Total RNA was extracted from patient plasma using the QiAmp viral RNA mini kit (Qiagen), and for oral samples using the Zymo Research Direct-zol RNA MiniPrep Kit (Zymo Research, ZR R2052) according to the manufacturer’s instructions.

Sequencing libraries were prepared from all samples. Briefly, DNA was removed by Turbo DNase treatment (Thermo Fisher Scientific), human RNA was depleted in a sample with more than one million reads of rRNA quantified by 18S qPCR. cDNA was synthesized with random priming and SuperScript IV (Thermo Fisher Scientific) and sequencing libraries were prepared using the DNA Prep Library Prep Kit (Illumina) as previously described (78). A total of 206 libraries were made at CIGASS and sent for sequencing on either the Illumina NovaSeq and NextSeq instrument at the Broad Institute, with paired end 100nt sequencing.

### Pan-viral hybrid capture

A total of 25 samples were selected for Hybrid capture. These 25 samples showed viruses in Kraken, Sourmash or both on the original run. For samples selected for viral enrichment, total RNA was extracted from a new sample aliquot, using the kits and methods described above, at Brown University. RNA quality and yield were assessed by 18S RT-qPCR using Applied Biosystems Power SYBR Green RNA-to-Ct *1-Step* kit (Catalog no: 4389986). Dnase treatment was performed using Invitrogen Turbo DNA-free kit (catalog no: AM1907) according to the manufacturer’s instructions. Libraries were prepared using TWIST whole transcriptome sequencing reagents with rRNA Depletion and were assessed using Agilent tapestation. Qubit High Sensitivity dsDNA kit was used to quantify libraries which were then normalized and pooled into 4 pools, 2 pools of oral samples and 2 pools of plasma samples. Hybrid capture was performed on the sample pools using TWIST Bioscience’s comprehensive viral research panel and Standard Hybridization kit from TWIST following the manufacturer’s instructions. The capture enriched pools were sequenced on an Illumina NovaSeq with 150bp paired end sequencing.

### Complete 16S Sequencing and Taxonomic profiling

The full 16S rRNA (V1-V9) was amplified with the 27 F/1492 R (27 F: 5’-AGAGTTTGATCMTGGCTCAG-3’) and 1492 R: 5’-TACGGYTACCTTGTTAYGACTT-3’) primer (79). DNA was amplified using conditions described in the protocol (80) with 32 cycles using the Thermo Scientific Phusion High-Fidelity DNA. PCR products were purified with 0.7 X AMPure XP beads (Beckman Coulter, A63880). DNA libraries were prepared using the Ligation sequencing amplicons – Native Barcoding Kit 96 V14 (SQK-NBD114.96) protocol. For Oxford Nanopore sequencing. Libraries were purified with a 1:1 ratio of AMPure XP beads and loaded into the Oxford Nanopore, FLO-MIN114, R10.4.1 chemistry. Generated fastq were constructed and filter targeting 1.2-1.8 kb using SeqKit2 (81). We employed epi2me-labs/wf-16s nextflow workflow (v1.6.0) (82) with Kraken2 as classifier with a confidence of 0.001 for taxonomic classification, abundance estimation and alpha diversity estimation of 16S rRNA samples.

### Metranscriptomic Taxonomic profiling

We used the KneadData pipeline (29) for adapter trimming, quality filtering, and removal of human host reads. Taxonomic profiling on cleaned data was performed using three different tools Kraken2 (32), Sourmash v4.7.0 (33) and MetaPhlan v4.2.4 (38). For Kraken2 we use the PlusPF database (build 20221209) and reads were classified in paired-end mode with a confidence threshold of 0.05. For sourmash, signatures were generated using k-mer sketches k = 31, a scaling factor of 100, and abundance tracking enabled. The GenBank viral, bacterial, fungal, and protozoan genomes (release March 2022) were used with a minimum threshold of 0 bp. Taxonomic assignments for gather hits were obtained using sourmash taxonomy annotate, leveraging GenBank lineage metadata. For MetaPhlan we estimate abundance with the database vJan25.

Kraken2 outputs were converted into BIOM format using kraken_biom v1.2 (83), sourmash taxonomic annotate table were converted into phyloseq objects using the sourmashconsumr package (84) and MetaPhlan abundance file were also used to create phyloseq objects. The R package phyloseq (85) were used for abundance plotting and diversity estimation. Shannon diversity and Bray-Curtis were only estimated using Kraken2 data. Alpha-diversity metrics (Shannon Index) were calculated at the genus level for oral sample, locality, and gender using Kruskal–Wallis tests. Associations between continuous metadata (age, temperature) and diversity were evaluated by Spearman’s rank correlation. Bacteria species abundances over body temperature were modeled using LOESS (LOcally Estimated Scatterplot Smoothing) regression. Beta diversity was assessed using Bray–Curtis dissimilarity and visualized via PCoA with 95% confidence ellipses. Group differences were evaluated using PERMANOVA.

Contaminant removal was performed using both data-driven and literature-based strategies. The prevalence method implemented in the decontam package (30) (threshold = 0.5) was applied at the genus level to identify taxa enriched in negative controls. Additionally, taxa belonging to phyla known to represent common laboratory, reagent, or low-biomass contaminants were removed based on published contaminant lists (86, 87), we also removed all taxa contaminants in biological samples by excluding taxa whose abundance was less than twice the maximum observed in the negative control (NTC) samples.

### Detection and classification of viruses in metatranscriptomic data

For viral detection we used the Kraken2 and Sourmash assignments described above but with more permissive cut-offs to reflect the low abundance and genetic diversity of viral infections. Specifically, we considered a virus as present in a sample if it was detected by either Sourmash or Kraken2 at the taxonomic level of “genus” with a cut-off of 1 signature or 5 reads, respectively. We confirmed suspicious hits by read alignment to a reference genome: in one sample (LNG027) Kraken2 identified 12 reads matching the Ebola virus, however alignment to the reference genome (NC_002549.1) using minimap2 resulted in 0 aligned reads, so this result was considered a false positive. We used the same cut-offs for analyzing metatranscriptomic and capture-enriched data.

Following SPAdes(88) assembly, viral sequences were identified via DIAMOND v2.0.15 (89) and validated using the taxize (90) R package, applying stringency filters of ≥500 bp contig length and 20–80% amino acid identity.

### Viral genome assembly and phylogenetic analysis

Genome assembly for viruses was done using Viral-ngs pipeline from BROAD Institute (https://viral-pipelines.readthedocs.io/en/latest/). Genomes of known viruses were assembled using reference-based genome assembly from viral-NGS pipeline with their default settings.

To investigate the relationship among our collected Redondoviridae samples, we retrieved 139 Redondoviridae samples from NCBI Virus. Sequences were then filtered to include complete genomes, which is defined as sequences with greater than 3,000 nucleotides (54). This results in a total of 102 sequences. We further excluded 7 sequences that showed evidence of potential misalignment or sequencing errors, giving a final dataset of 95 sequences. We performed multiple sequence alignment of the 100 sequences using MAFFT v7.505 (91) with default global alignment. Aligned sequences were then trimmed using trimAl v1.4. rev15 (92) to remove poorly aligned regions. Maximum-likelihood phylogenetic trees were constructed using IQ-TREE v2.1.4 with 1000 bootstrap replicates. Trees were visualized using the package ggtree v3.16.3 in R 4.5.1. Sample tips were colored by country of origin, but because country information was not available for all NCBI entries, we consulted the original publications to obtain missing metadata. As our samples yielded complete coverage for only the capsid gene, we performed a separate phylogenetic analysis specific for the capsid. For this, we re-aligned our sequences to a vientovirus reference genome using MAFFT and then trimmed to only include the capsid gene region. Phylogenetic tree reconstruction followed the same pipeline described above.

### Metagenomic assembly, AMR and Virulence factor detection

Individual assembly on 150 samples were made using MEGAHIT v1.2.9 with minimum contig length of 500 base pairs. To identify antimicrobial resistance genes, assembled contig files from each sample were screened using the Resistance Gene Identifier (RGI) tool. RGI was run individually on each contig file with parameters set for local execution, using Prodigal’s algorithms for low quality/coverage assemblies using the Comprehensive Antibiotic Resistance Database (CARD). Gene with Strict and Perfect matches were kept for analysis. Contigs ID classified as ARG were subsequently identified at the species level using local BLASTn. BLAST hits were retained if the e-value ≤ 1×10□□, percent identity ≥ 80%, and query coverage ≥ 80%. To identify if contigs classified as ARGs were from plasmid, chromosome or viral origin we used the geNomad end-to-end pipeline and the MEGAHIT assembly contigs fasta file as input using the default parameters and database. To identify virulence factors genes, assembled contig files from each sample were screened using ABRicate with the VFDB (Virulence Factor Database). ABRicate was executed on each contig file individually. These results were subsequently used for downstream network analyses. Species with VF were identified using NCBI BLAST with the same parameters used for ARGs contigs species identification.

## Contributions

K.J.S., P.C.S., D.N., M.F.P. and A.S.B. conceived the study and secured funding. I.M.N., A.M.M., Y.D., M.T., J.F.G coordinated participant enrollment and sample collection. A.G., T.N., M.S., R.T generated sequencing data. M.S., R.T., D.F, Z.C.L, K.L.B, and K.J.S. analyzed the data. M.S., T.N, R.T, A.G., Z.C.L, B.N., K.L.B, D.F., M.T., I.M.N, Y.D., A.M.M., J.F.G., K.C.D., A.B.D., M.N., A.S.B., M.F.P., P.C.S., D.N. and K.J.S. drafted and revised the manuscript with input from all authors. All authors approved the manuscript before submission.

## Acknowledgments

We would like to express gratitude to the community of Tambacounda and Linguere, and to all the healthcare workers at the clinics including medical directors, doctors, nurses, and clinic staff for their participation and support in the collection of samples required for this analysis. We also thank Dr. Peter Belenkey and Dr. Stephen Costa for sharing materials and expertise for the complete 16S sequencing, and Dr. Karthikeyani Chellappa for providing positive control samples for 16S rRNA protocol validation.

## Data availability

Raw sequencing reads and assembled genomes will be deposited in the Sequence Read Archive and NCBI GenBank prior to publication. Raw counts for metagenomic classification and the code used for analysis are available here: https://github.com/lab-SIDL/Oral-and-plasma-microbiome-in-the-context-of-acute-febrile-illness

---

**Supplementary Figure 1 :**
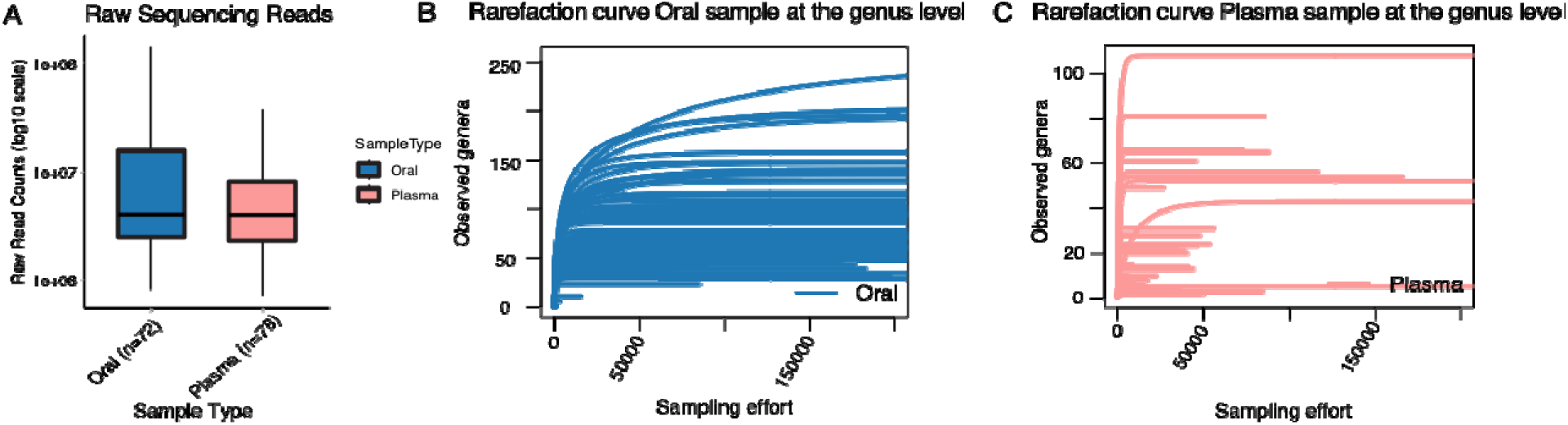
A) Sequencing depth, B) Rarefaction curve oral sample at the genus level, C) Rarefaction curve plasma sample at the genus level.

**Supplementary Figure 2:**
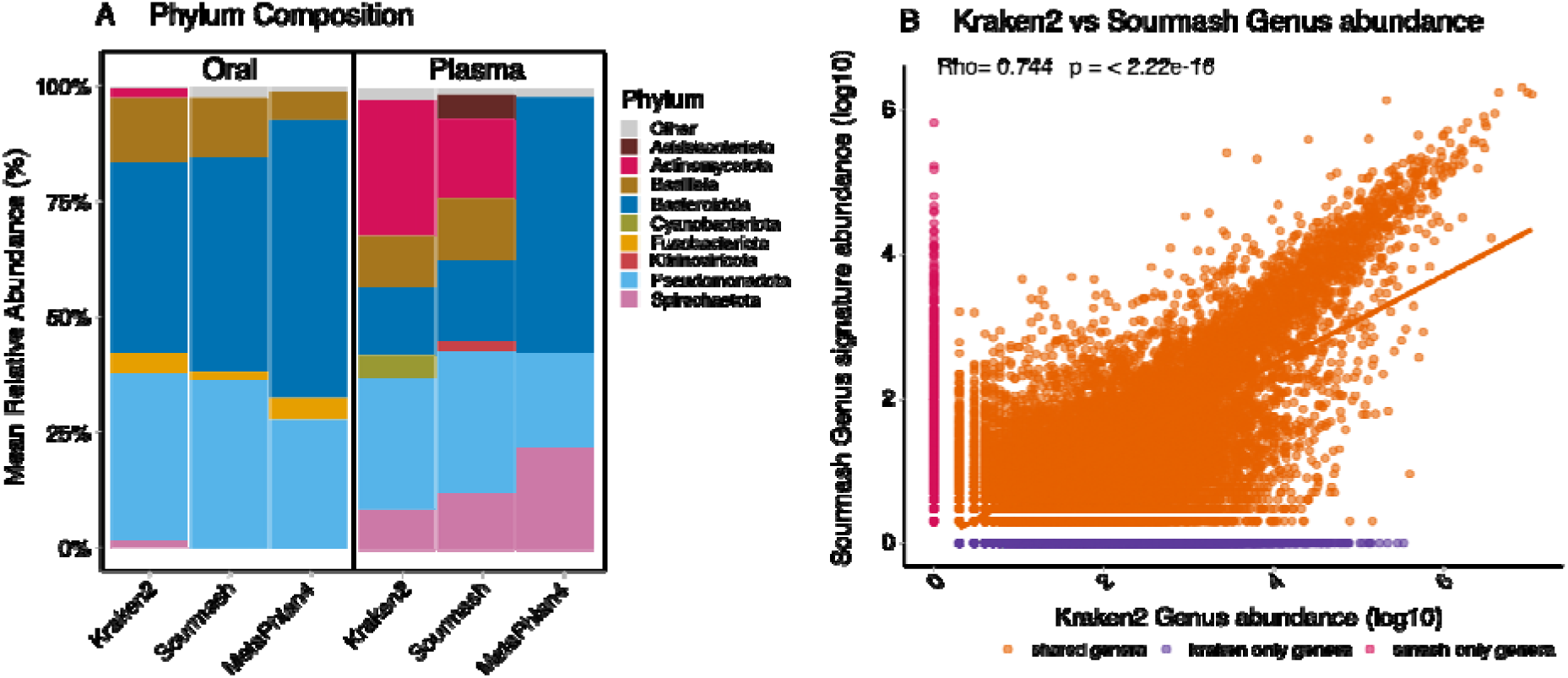
Phylum mean relative abundance and correlation plots between Kraken2 and Sourmash.

**Supplementary Figure 3:**
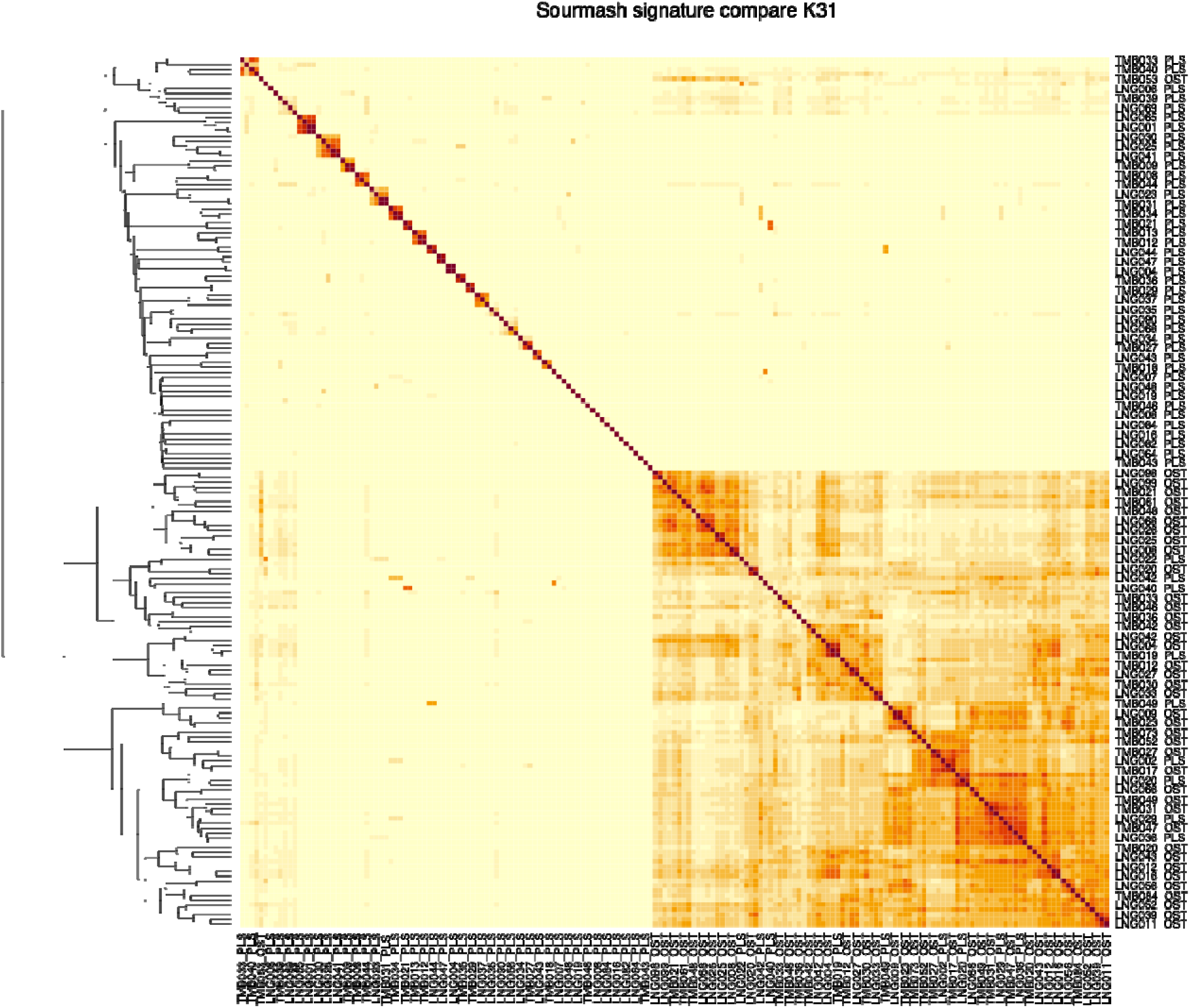
Sourmash signature comparison (k-mer = 31). The dark red indicates high-similarity between samples.

**Supplementary Figure 4:**
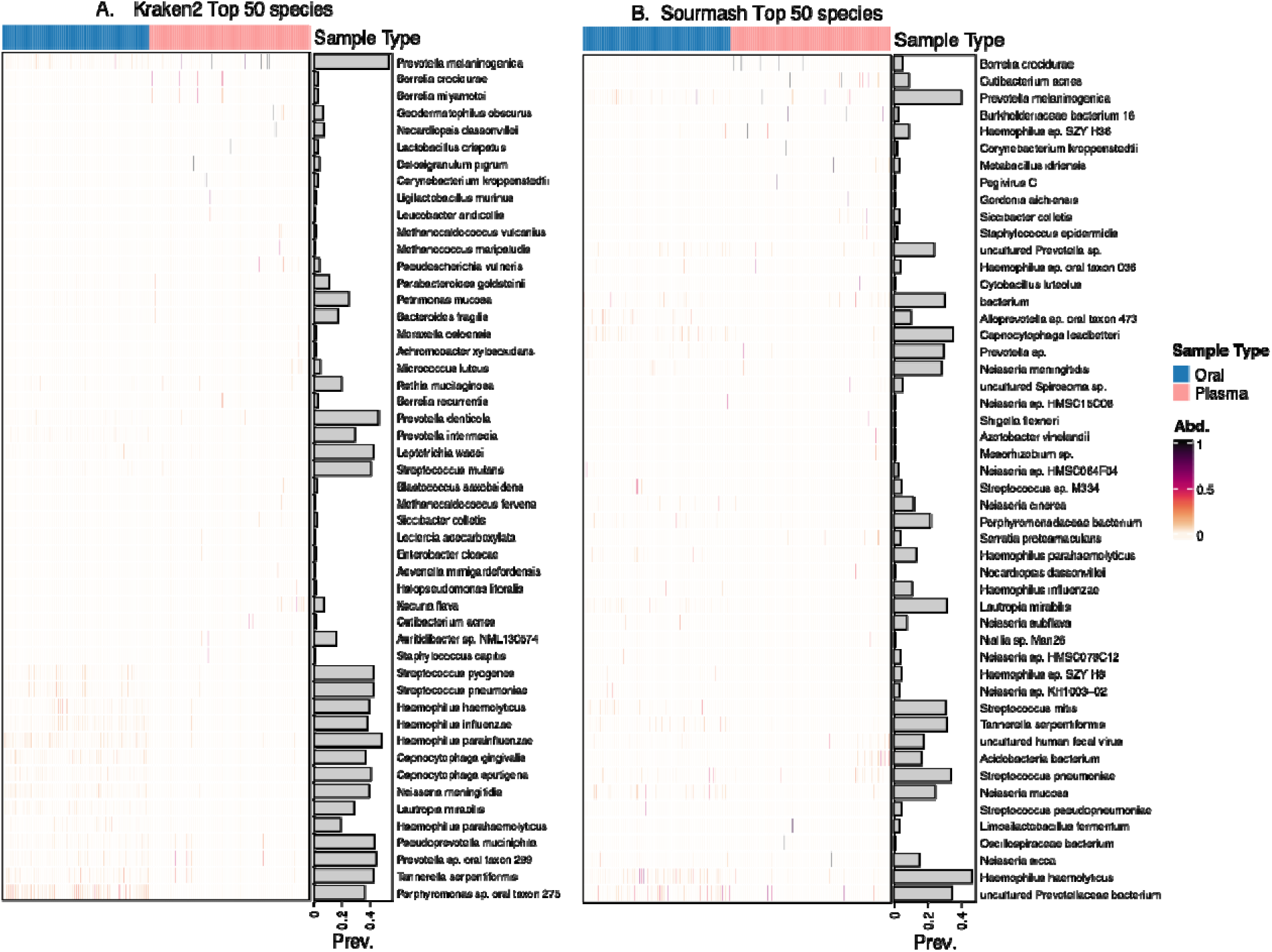
Top 50 species detected by kraken2 (A) and Sourmash (B)

**Supplementary Figure 5 :**
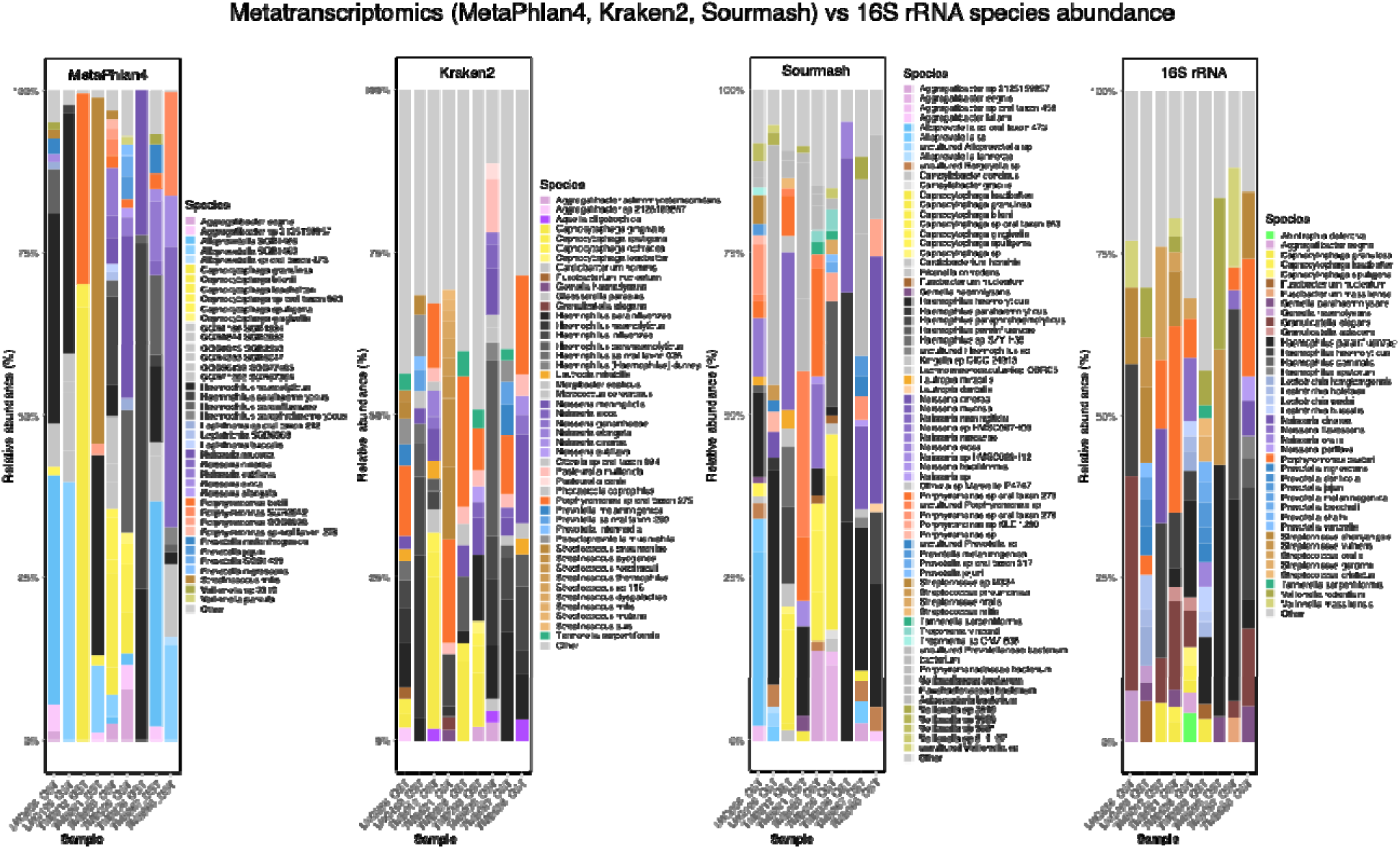
16S rRNA and metatranscriptomic species abundances

**Supplementary Figure 6 :**
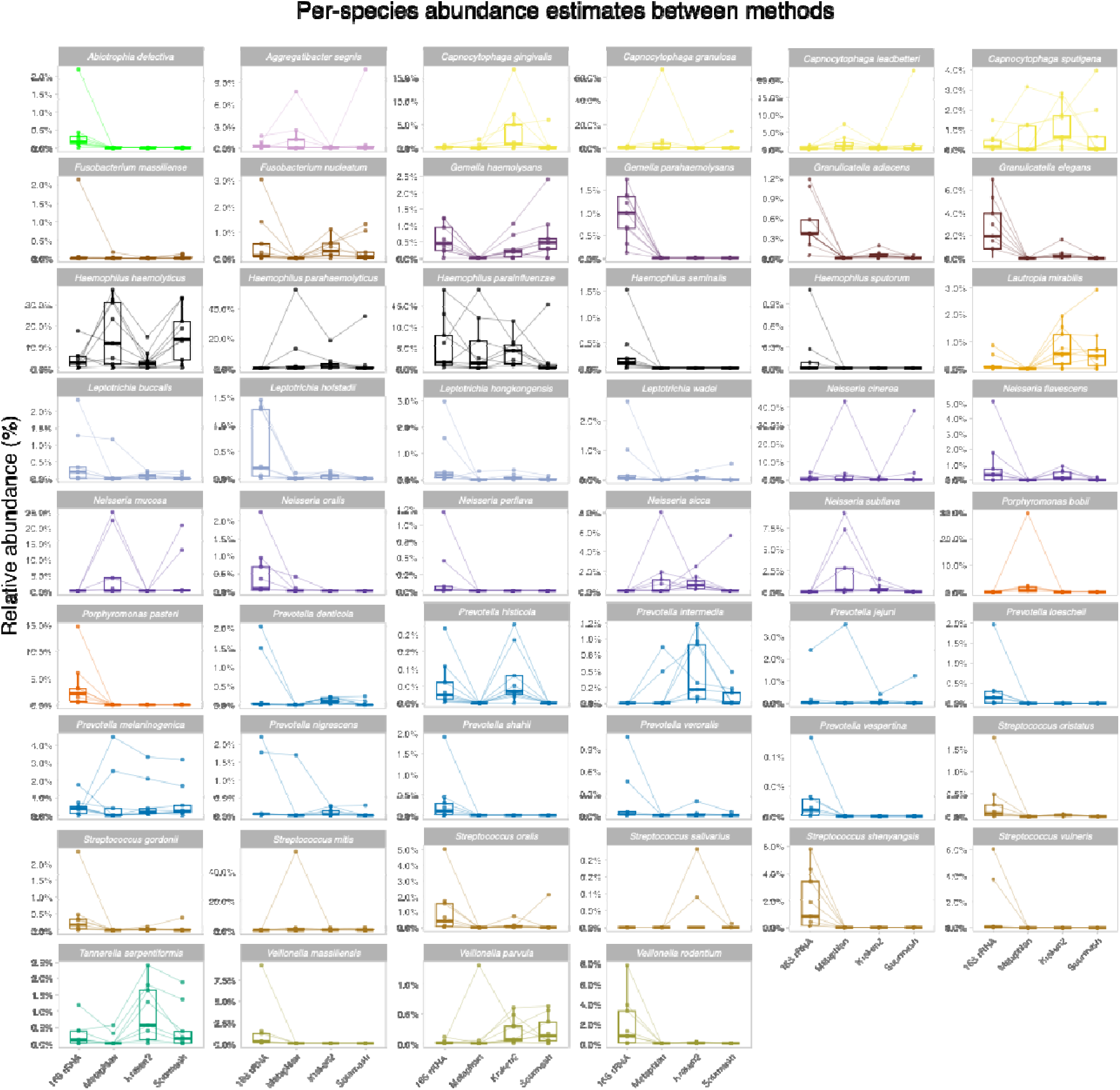
16S top species abundance compared to abundance found with metatranscriptomic analysis with kraken2, sourmash and Metaphlan4.

**Supplementary Figure 7 :**
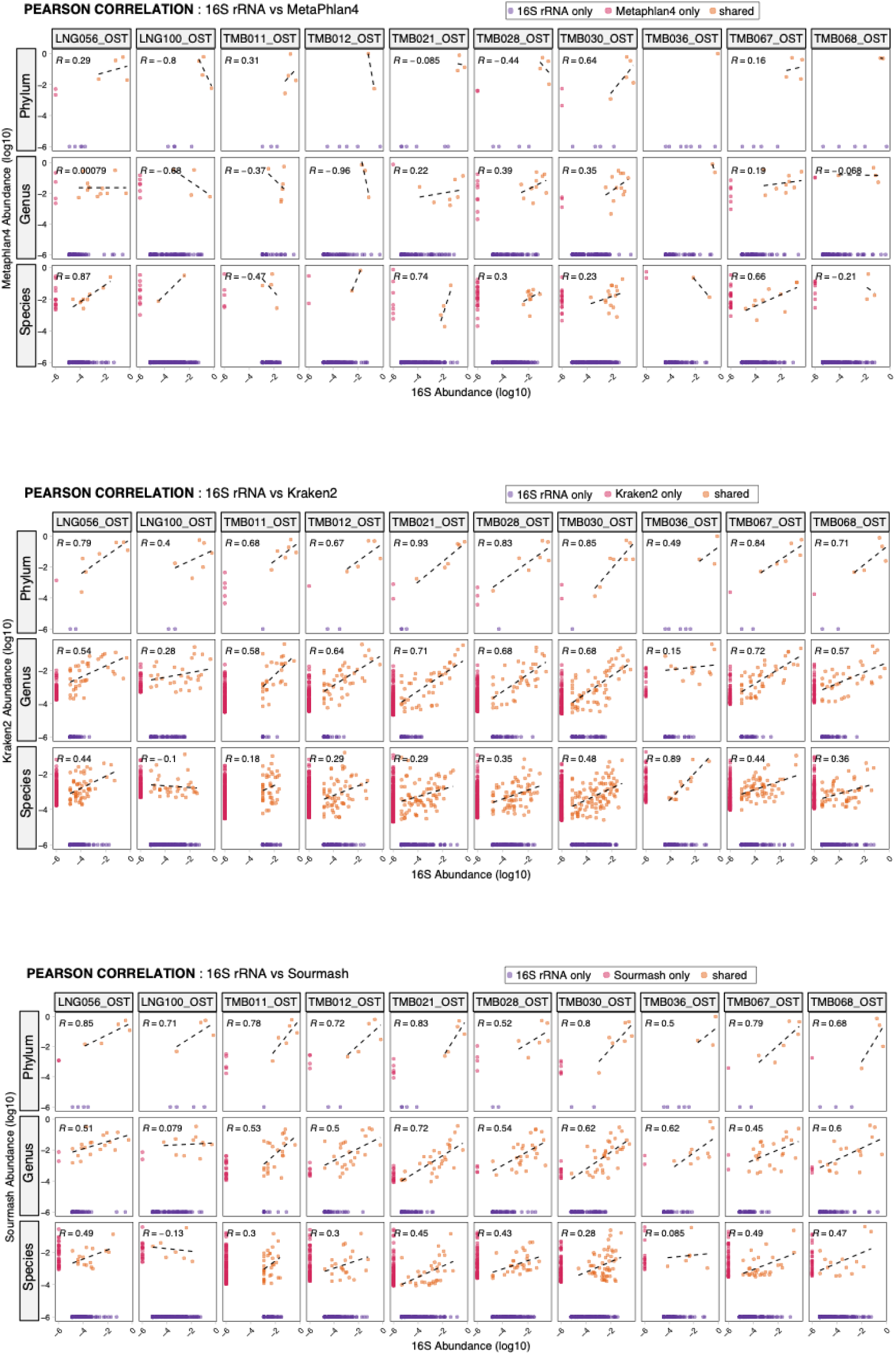
Pearson correlation between 16S rRNA and Metatranscriptomic tools at the individual sample at the Phylum, Genus and Species level. Correlation analysis was performed only at the share taxon.

**Supplementary Figure 8:**
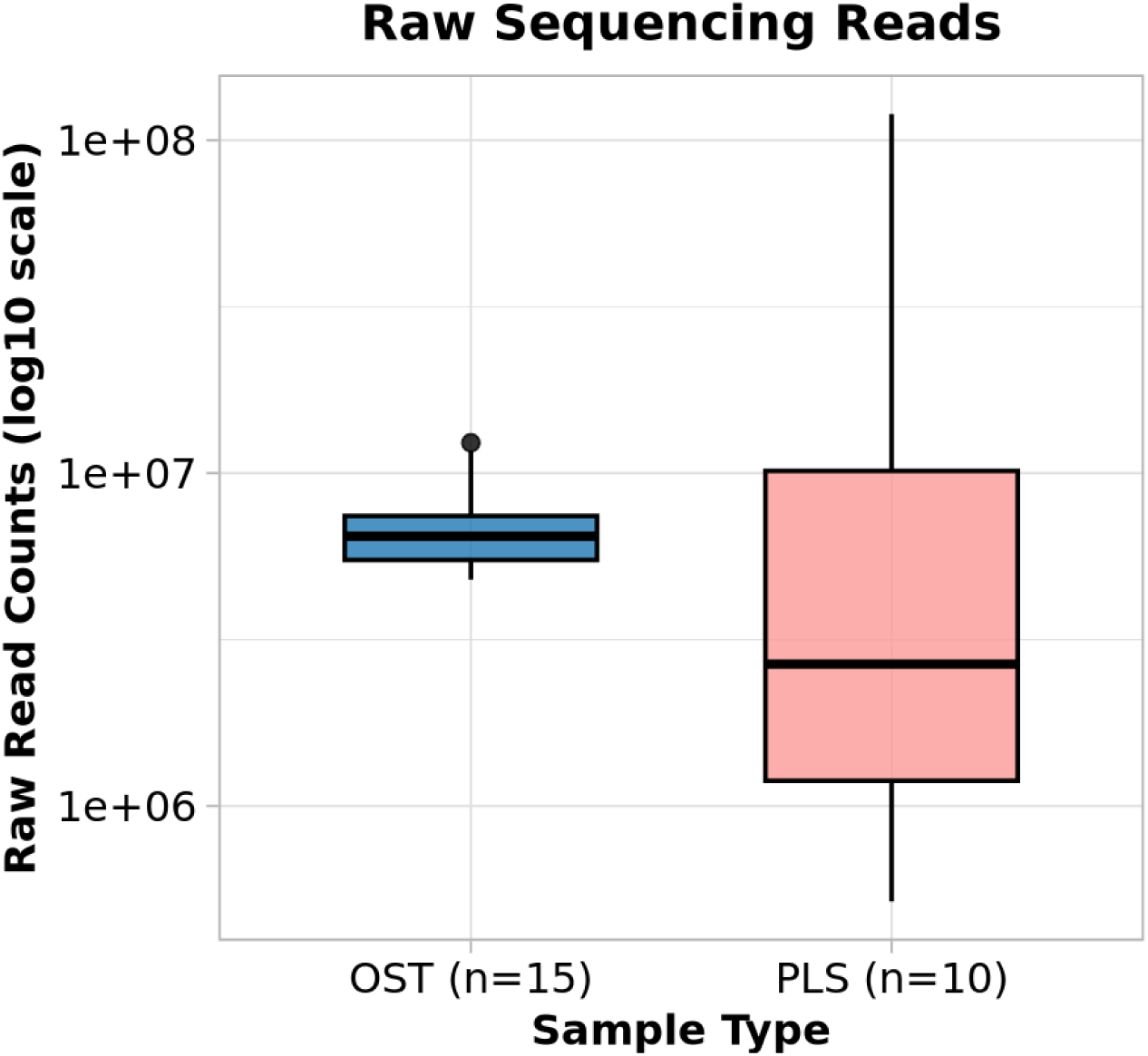
Hybrid capture sequencing depth. Boxplot showing raw read depth for oral (OST, N=15) and Plasma (PLS, N=10) samples after hybrid capture enrichment with pan-viral enrichment probes.

**Supplementary Figure 9:**
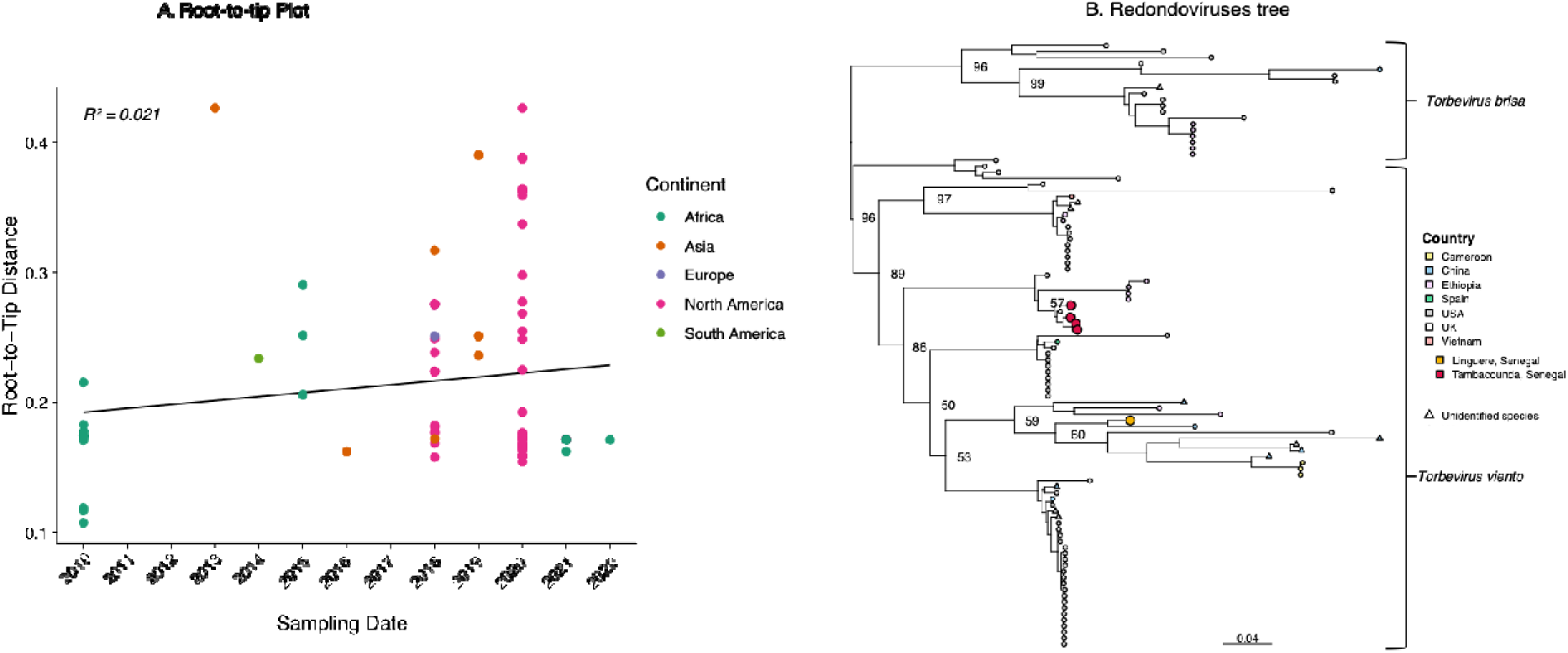
Phylogenetic investigation of redondoviruses. A. Root-to-tip plot of all vientovirus samples. B. maximum likelihood tree of the viral Cap gene.

**Supplementary Figure 10:**
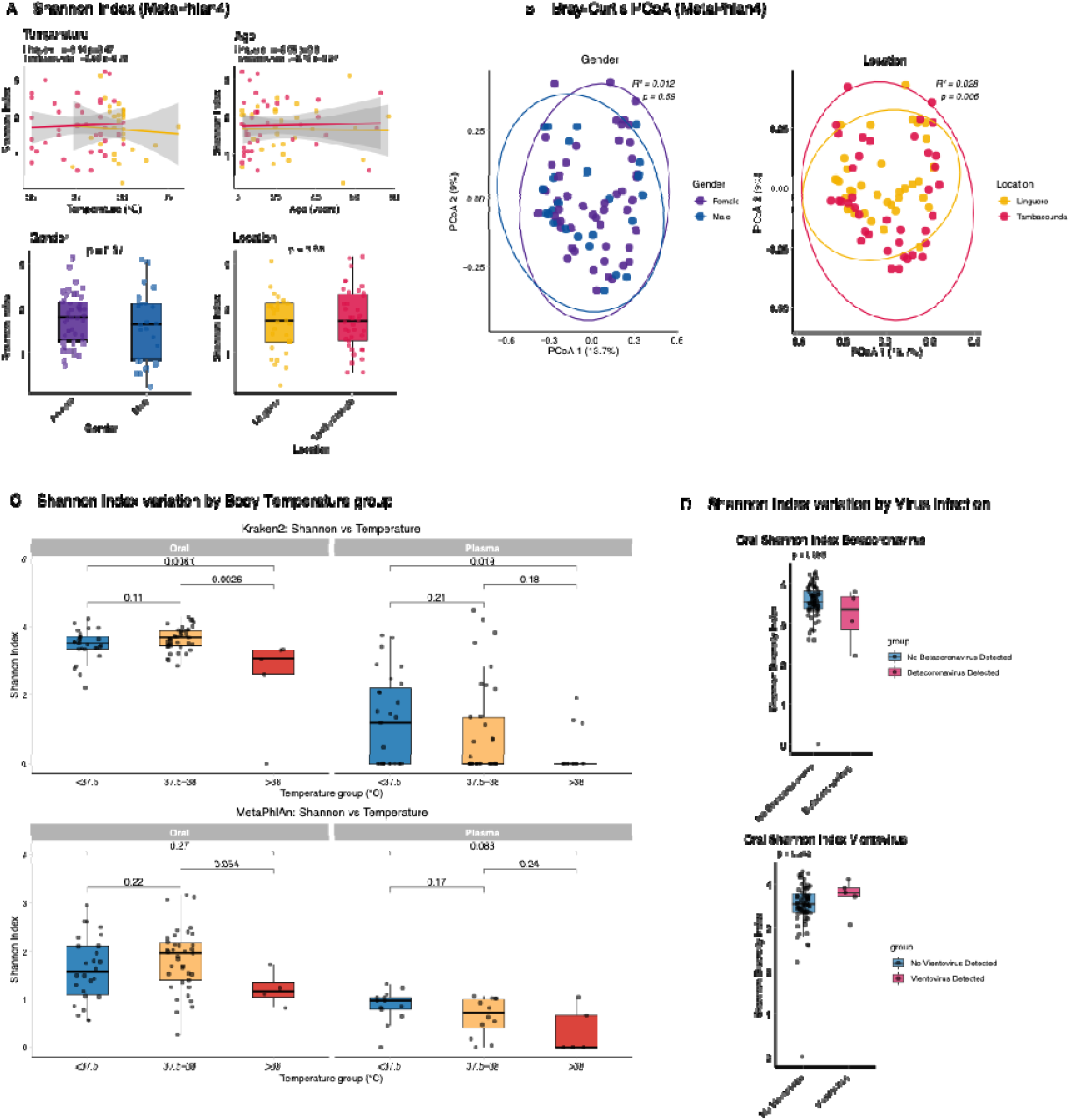
A) Oral sample Shannon Index with MetaPhlan for Temperature, Age, Gender and Location, B) Oral sample Bray-Curtis principal coordinate analysis (PCoA) plot depicting differences in microbial composition for Gender and Location with MetaPhlan, C) boxplots of Shannon Index for different body temperature groupings using Kraken2 and MetaPhlan abundance estimates for plasma and oral sample, D) Shannon index comparison between group infected with SARS-CoV-2 and Vientovirus and group with no viral infection.

**Supplementary Figure 11:**
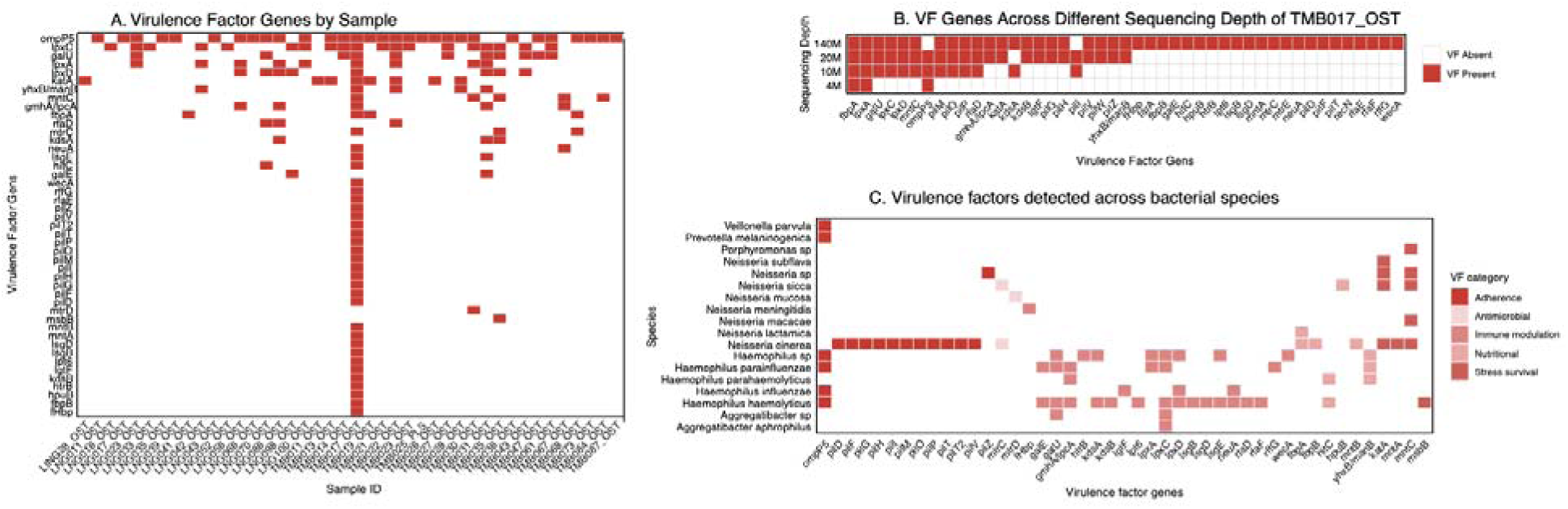
A) Virulence factor (VF), B) VF detected in sample TMB017_OST across different sequencing depth C) Bacterial Species with VF Genes.

**Supplementary Table 1:** Contaminant list for filtering.

| **Taxonomic Group** | **Genera** | **Source** |
| --- | --- | --- |
| **Proteobacteria** | *Aquabacterium, Bradyrhizobium, Brevundimonas, Hoeflea, Methylobacterium, Phyllobacterium, Rhizobium, Acidovorax, Azoarcus, Burkholderia, Comamonas, Cupriavidus, Delftia, Duganella, Herbaspirillum, Janthinobacterium, Leptothrix, Sphingomonas, Limnobacter, Massilia, Methyloversatilis, Polaromonas, Ralstonia, Undibacterium, Acinetobacter, Escherichia, Nevskia, Pseudomonas, Stenotrophomonas, Xanthomonas* | Salter et al. (2014) |
| **Actinomycetota** | *Propionibacterium, Actinomyces, Micromonospora* | Salter et al. (2014);<br>Weyrich et al. (2019) |
| **Bacillota** | *Bacillus, Exiguobacterium, Jeotgalicoccus* | Salter et al. (2014);<br>Weyrich et al. (2019) |
| **Bacteroidota** | *Flavobacterium* | Salter et al. (2014) |
| **Other** | *Enhydrobacter, Stutzerimonas, Rheinheimera, Alcaligenes, Methanotorris* | Weyrich et al. (2019) |

**Supplementary Table 2:** Known pathogens participant demographic characteristics.

| **Bacteria** | **Viruses** | **Age range (years)** | **Sex** | **Temperature (°C)** |
| --- | --- | --- | --- | --- |
| *Borrelia crocidurae* |  | 11–15 | Female | 39.3 |
| *Borrelia crocidurae* |  | 1–5 | Female | 39 |
| *Borrelia* |  | 26–30 | Female | 39 |
| *Borrelia crocidurae* |  | 56–60 | Male | 39.8 |
| *Borrelia crocidurae* | Lymphocryptovirus | 1–5 | Female | 37.3 |
| *Borrelia* |  | 21–25 | Female | 37.5 |
| *Borrelia recurrentis* |  | 1–5 | Female | 38 |
| *Borrelia crocidurae* |  | 51–55 | Male | 38.5 |
|  | SARS-CoV-2 | 66–70 | Female | 37.8 |
| *Borrelia* | Mastodonovirus | 6–10 | Female | 38 |
|  | SARS-CoV-2 | 26–30 | Female | 37.9 |
|  | SARS-CoV-2 | 6–10 | Male | 37.7 |
| *Borrelia crocidurae* |  | 16–20 | Male | 37.8 |
| *Borrelia crocidurae* |  | 16–20 | Male | 36.6 |
| *Borrelia* |  | 46–50 | Female | 36 |
|  | Lymphocryptovirus | 1–5 | Female | 37 |
|  | Lymphocryptovirus | 1–5 | Female | 37.6 |
| *Borrelia crocidurae* |  | 71–75 | Female | 37 |
|  | Simplexe | 1–5 | Female | 37.6 |
|  | SARS-CoV-2 | 66–70 | Male | 37.3 |
|  | Lymphocryptovirus | 41–45 | Female | 37.5 |
| *Borrelia* |  | 6–10 | Female | 36 |
|  | SARS-CoV-2 | 1–5 | Male | 36.6 |

